# Vaginal Microbiota Transplantation (VMT) for treatment of vaginal dysbiosis without the use of antibiotics – A Double-Blinded Randomized Controlled Trial in healthy women with vaginal dysbiosis

**DOI:** 10.1101/2024.06.28.24309465

**Authors:** Tine Wrønding, Kilian Vomstein, Kevin DeLong, Agnete Troen Lundgaard, Sarah Mollerup, Brynjulf Mortensen, Elleke F. Bosma, Ann Marie Hellerung, Emilie Vester Engel, Klara Dortea Wiil, Julie Elm Heintz, Sofie Ingdam Halkjær, Luisa W Hugerth, Tanja Schlaikjær Hartwig, Andreas Munk Petersen, Anne Bloch Thomsen, David Westergaard, Nina La Cour Freiesleben, Henrik Westh, Johan E.T. van Hylckama Vlieg, Laura M. Ensign, Henriette Svarre Nielsen

**Affiliations:** Department of Obstetrics and Gynecology, The Fertility Clinic, Copenhagen University Hospital, Hvidovre, Denmark; Freya Biosciences Aps, Fruebjergvej 3, 2100 Copenhagen, Denmark; Department of Clinical Microbiology, Copenhagen University Hospital, Hvidovre, Denmark; Gastrounit, Medical Division, Copenhagen University Hospital Hvidovre, Hvidovre, Denmark; Department of Clinical Medicine, Faculty of Health and Medical Sciences, University of Copenhagen, Copenhagen, Denmark; Department of Medical Biochemistry and Microbiology, Science for Life Laboratory, Uppsala University, Uppsala, Sweden; Center for Nanomedicine at the Wilmer Eye Institute, Johns Hopkins University School of Medicine, Baltimore, MD 21231, USA; Department of Chemical & Biomolecular Engineering, Johns Hopkins University, Baltimore, MD 21218, USA; Department of Ophthalmology, Wilmer Eye Institute, Johns Hopkins University School of Medicine, Baltimore, MD 21287, USA; Department of Pharmacology and Molecular Sciences, Johns Hopkins University School of Medicine, Baltimore, MD 21287, USA; Departments of Gynecology and Obstetrics, Infectious Diseases, and Oncology, Johns Hopkins University School of Medicine, Baltimore, MD 21287, USA; Department of Biomedical Engineering, Johns Hopkins University, Baltimore, MD 21218, USA

**Author notes:** Corresponding author: Henriette Svarre Nielsen.

**Keywords:** Vaginal dysbiosis, Vaginal Microbiota Transplantation, Antibiotic treatment, Antiseptic pretreatment, Immunological changes, Recurrent BV, Randomized Controlled Trial

## Abstract

Here we describe the first double-blinded, randomized, placebo-controlled trial (RCT) on vaginal microbiota transplantation (VMT) without antibiotics in women with *both* symptomatic and asymptomatic vaginal dysbiosis. Forty-nine women were randomly assigned to VMT or placebo. The trial did not show a significant conversion to our predefined *Lactobacillus*-dominated microbiome. However, in participants not initially converting, antiseptic pretreatment before a subsequent VMT led to a 50% conversion rate, associated with an anti-inflammatory shift in gene expression. Metagenomic sequencing and strain-level genetic analysis confirmed donor engraftment in five of 10 women who showed microbiome conversion. Extensive exploration of the microbiome, immune response and metadata revealed differences in baseline energy metabolism in participants who later experienced donor engraftment. Treatments for vaginal dysbiosis are urgently needed and given that VMT can lead to donor engraftment and change the vaginal immune profile, future studies should focus on optimizing this treatment for various women’s health diseases.

## Introduction

A vaginal microbiome dominated by lactobacilli (*L. crispatus, L. gasseri, L. iners*, or *L. jensenii*) is associated with reproductive tract health (1–3). The production of lactic acid by these bacteria lowers the vaginal pH, typically to values below 4.5 (4). A more diverse vaginal microbiome dominated by other bacteria such as *Gardnerella* spp.*, Fannyhessea vaginae*, and *Prevotella* spp. is referred to as vaginal dysbiosis (1–3). Vaginal dysbiosis is estimated to be prevalent in 16-52% of women depending on study population (5) and geographic location (6,7) and is associated with a lower chance of conceiving in *in-vitro* fertilization (5), higher risk of euploid pregnancy loss (8), and obstetric complications such as premature rupture of membranes and preterm birth (9). Vaginal dysbiosis is lacking standardized diagnostic criteria and probiotics and antibiotics are being tested for their efficacy in treating vaginal dysbiosis (1,10). Bacterial vaginosis (BV) is characterized by an overgrowth of anaerobic bacteria, such as *Gardnerella vaginalis*, and a decrease in lactobacilli. BV is diagnosed using Amsel criteria (10) or Nugent score (11), and is typically treated with antibiotics. Key distinctions between BV and vaginal dysbiosis include the broader microbial imbalances in vaginal dysbiosis versus the specific microbial shifts in BV, with diagnostic and treatment protocols tailored accordingly (1,12,13). However, antibiotic treatment has some major drawbacks, including a recurrence rate of up to 60% within 12 months after treatment (12) due to high resistance rates to antibiotics (14–16). Also, orally administered metronidazole and clindamycin can disturb the gut microbiome (17), while vaginally applied antibiotics often result in subsequent vulvovaginal candidiasis (18).

To reduce recurrence rates after antibiotic treatment, a study from 2019 explored the use of vaginal microbiota transplants (VMT) sourced from healthy donors with a *Lactobacillus*-dominated vaginal microbiome. The study included five women with recurrent BV and four out of the five women showed clinical remission after up to three cycles of combined antibiotic and VMT interventions (19). In a case-study, our team successfully treated a woman with a complicated pregnancy history and severe symptomatic, *Gardnerella*-dominated vaginal dysbiosis with VMT without antibiotic pretreatment and observed donor strain engraftment (20). While these proof-of-concept studies provided promising results, the concept of VMT has not been tested in a randomized placebo-controlled trial (RCT) setting. Here we present a RCT testing VMT versus placebo in women with verified vaginal dysbiosis regardless of vaginal symptoms and test antiseptic pretreatment prior to VMT. In a companion study by Bosma and Mortensen et al., VMT is tested in asymptomatic women with vaginal dysbiosis using a different dosing regimen. Both studies aim to explore the immunological changes in relation to changes in the vaginal microbiota.

## Results

### Randomized Controlled Trial

Between June 1^st^ 2021 and June 2^nd^ 2022, we screened 302 healthy women of whom 51 (16.9%) met all eligibility criteria for participating in the study as a recipient (figure 1). Two of these women showed an eubiotic microbiome at the baseline visit and were excluded from further participation. Therefore, 49 women were randomly assigned to either VMT (n=37) or placebo (n=12) (saline water). The baseline characteristics for recipients are shown in table 1. The microbiome composition at all visits is shown in figure 2. The diversity in the vaginal microbiome in the VMT and the placebo group did not differ significantly at baseline. The most abundant taxon were *Gardnerella* spp., in 80-82% of the recipients (figure 2B). The primary outcome of the RCT trial was pre-defined as dysbiosis resolution (minimum of 70% total relative abundance of *Lactobacillus* spp. and a combined proportion of *Gardnerella* spp*., F. vaginae, and Prevotella* spp. of <10%) at any timepoint from first intervention to six month follow-up. After the first VMT, one out of 37 women (2.7%) in the VMT group changed her microbiome from dysbiotic to eubiotic. In the placebo group, two out of 12 women (15.4%) converted to eubiotic vaginal microbiome after the first intervention. We observed comparable rates of dysbiosis resolution following each dosing cycle. At the three-month follow-up, six months after the first intervention 11.4%, 4/35 in the VMT group and 37.3%, 3/11 in the placebo group had dysbiosis resolution. We found no significant difference between women receiving VMT versus placebo , with a hazard ratio (HR) of 0.71 (95% CI 0.18-2.76, p=0.64) (figure 3A). For the overall dysbiosis resolution counts see figure 3B.

**Figure 1:**
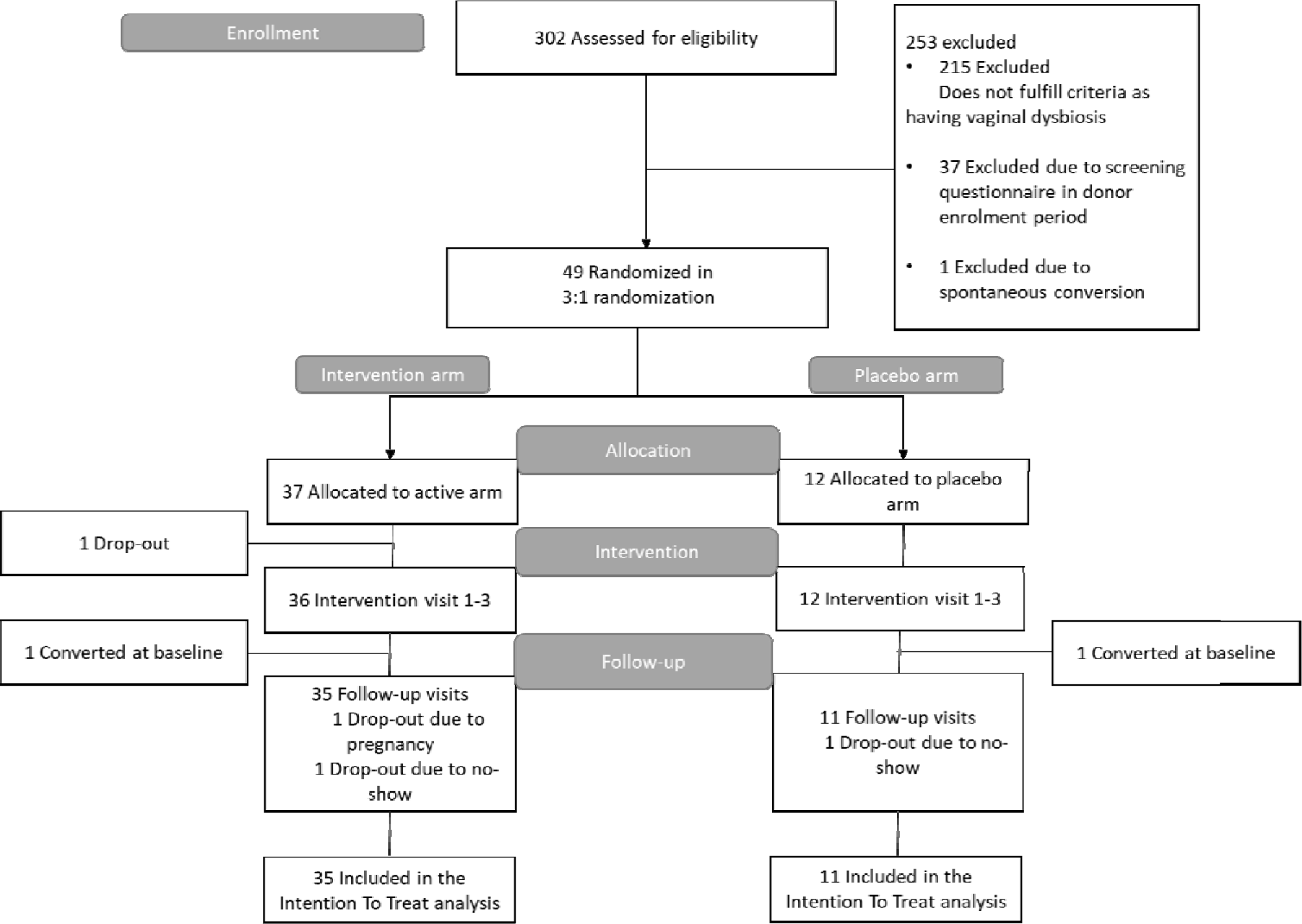
Trial flow diagram. Illustrated is a flow diagram of recipient participants within the VMT RCT study. Initially, potential participants underwent screening for microbiota composition and tests for various infections and conditions. Upon passing these screenings, they were randomly assigned in a 3:1 ratio to either the VMT or placebo intervention groups.

**Figure 2:**
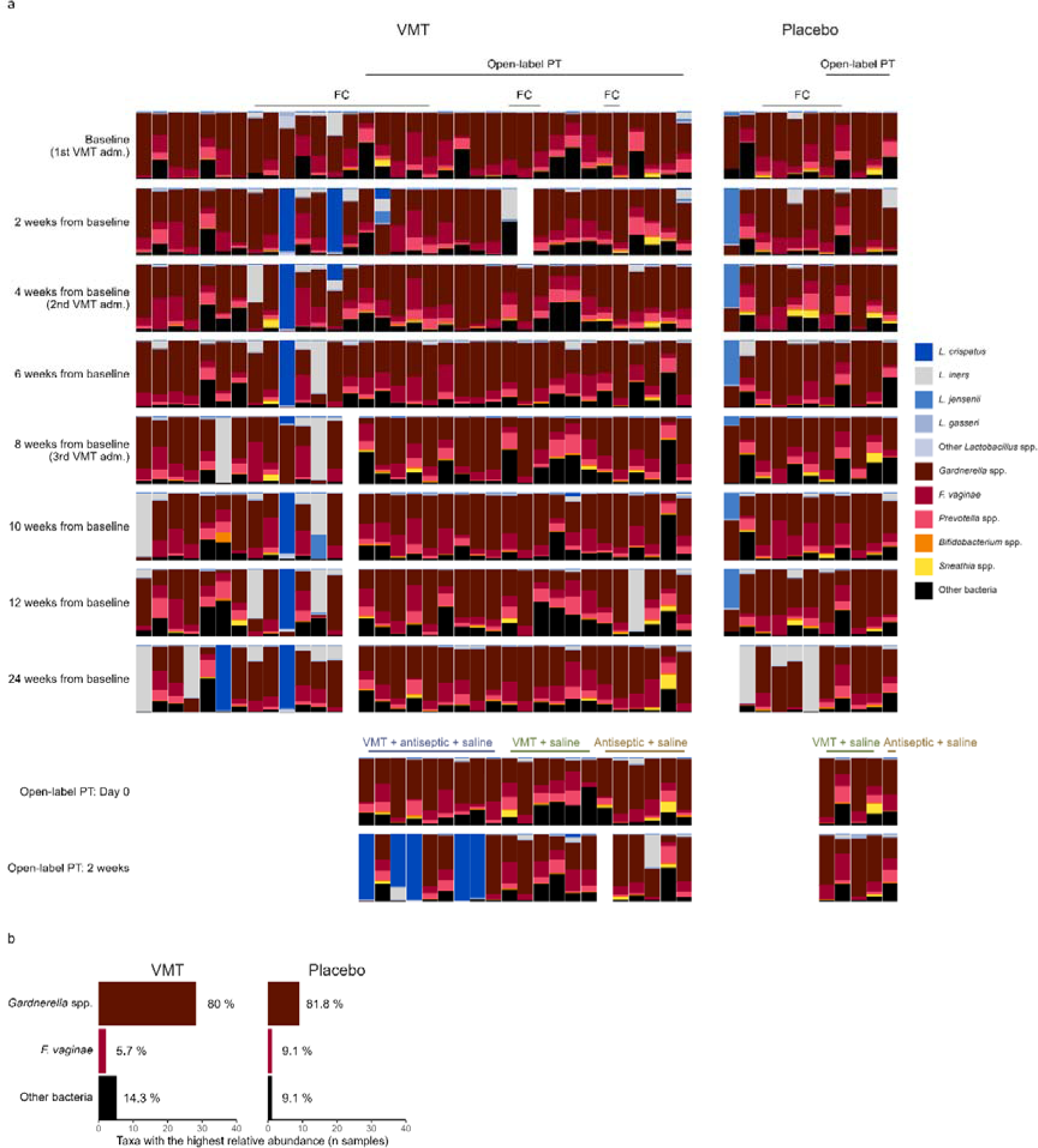
Bacterial composition in recipients. A: Stacked bar chart describing the microbiome composition of selected taxa at all visits. Flow cytometry (FC) and open-label pre-treatment (PT) participants indicated in top row for intervention (VMT) and placebo groups. B: Stacked bar chart showing the most dominant taxa for participants at baseline in the RCT study.

**Figure 3:**
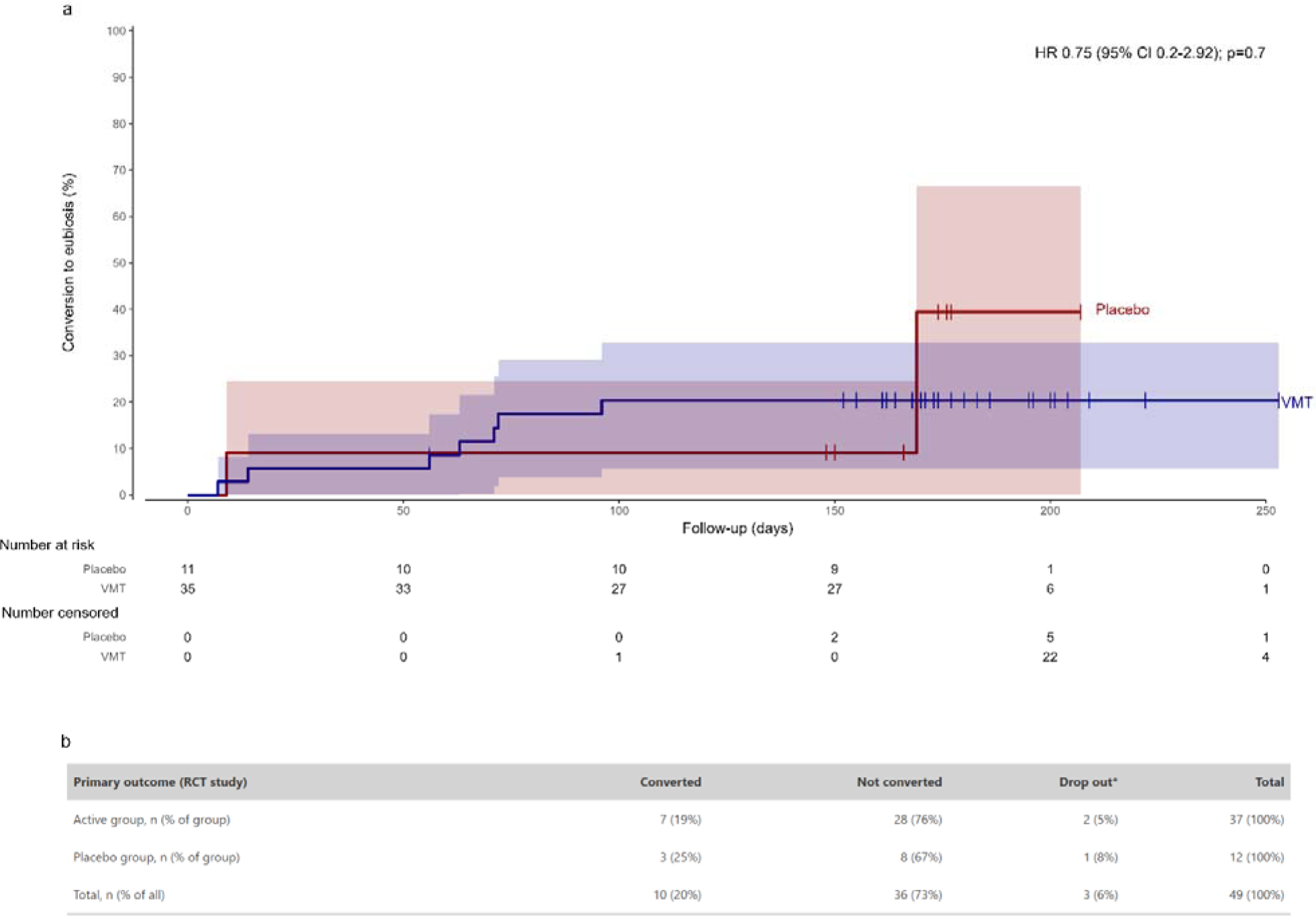
Treatment response rate in the RCT study. A: Kaplan-Meier estimates of conversion to eubiosis at any visit after the first dosing. Tick marks on the Kaplan-Meier curves represent individuals censured at the time of conversion to eubiosis or the last assessment. HR = Hazard Ratio. B: Percentages for primary outcome incl. dropouts.

**Table 1.**
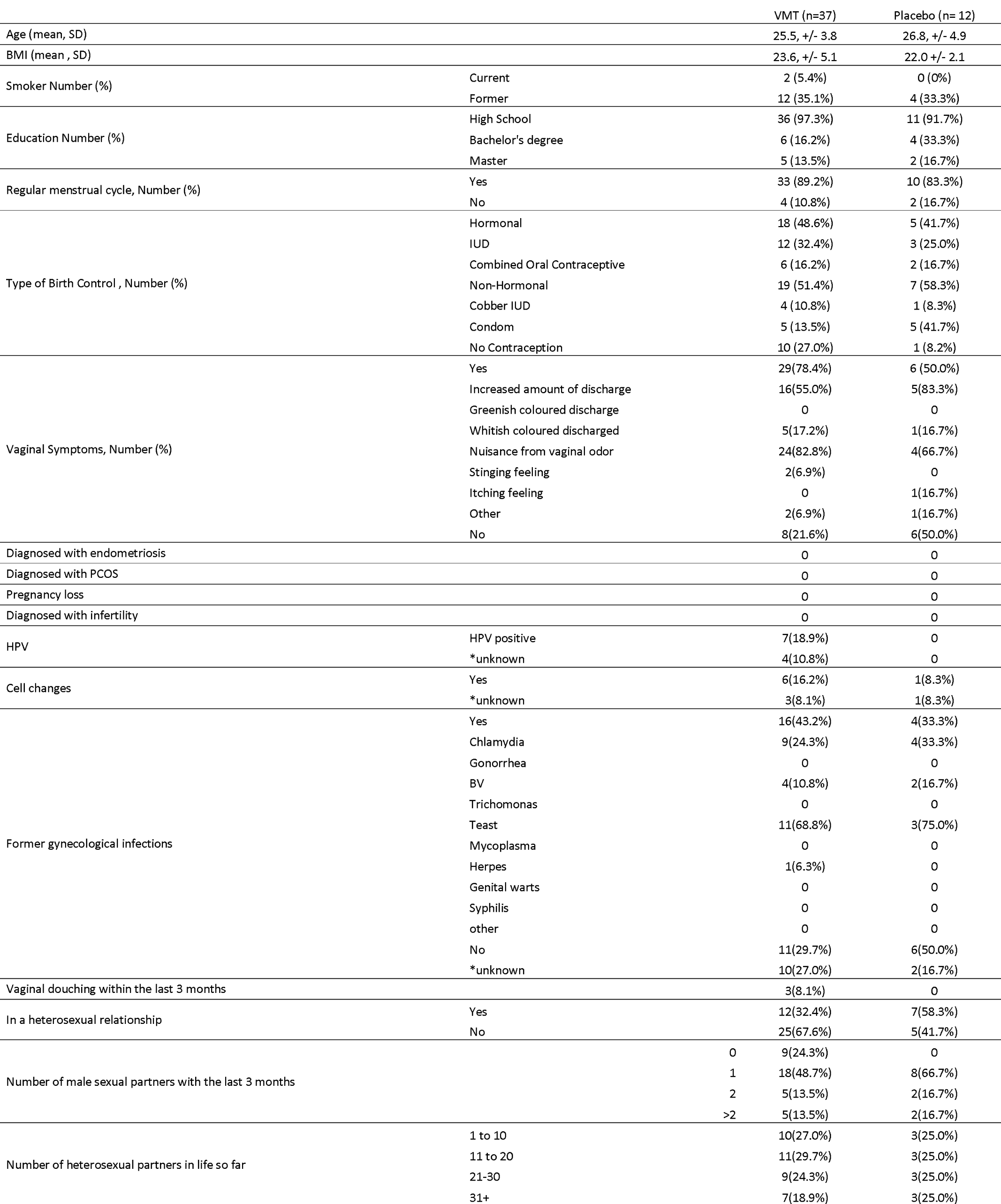
Demographic data on VMT and placebo-group. Data are n (%), mean (SD).

### Single nucleotide variant (SNV) analyses

SNV analyses were performed to evaluate if the converted microbiome in the VMT group was due to donor strain engraftment or spontaneous conversion. In two of the four women with dysbiosis resolution in the VMT group, we could identify donor strain engraftment (figure 4A and 4B). The two participants with donor strain engraftment also were the only ones of the converted who persistently showed a eubiotic microbiome composition 199 days after VMT (figure 4A and 4B).

**Figure 4:**
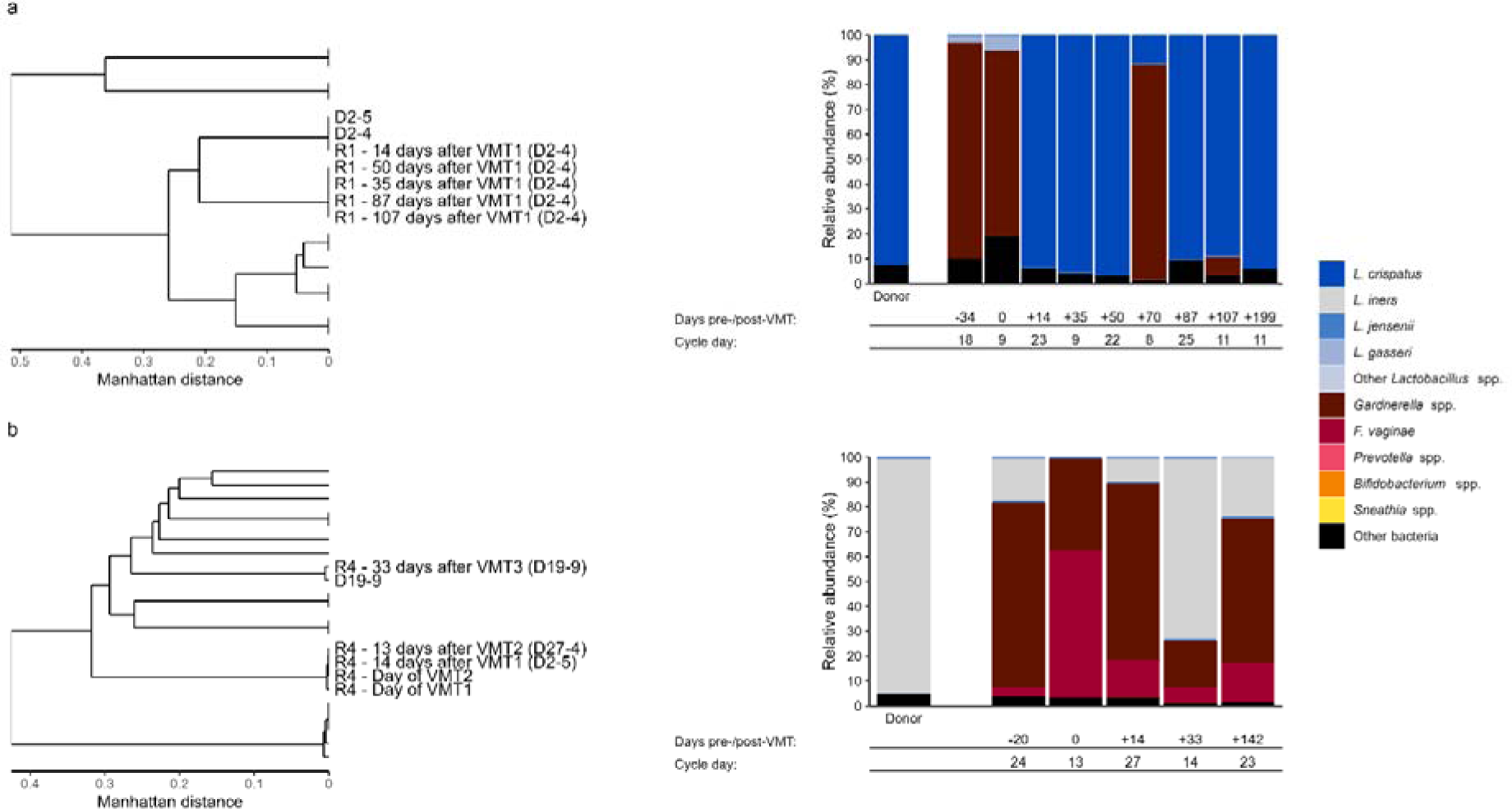
Single Nucleotide Variant (SNV) analysis of engraftment in RCT study To the left: Dendrograms of Manhattan distances between SNV profiles generated from metagenomic sequencing of CVS samples. The text in bold indicates samples from the donor-recipient pair with low genetic distance indicating successful donor engraftment. All other samples containing L. crispatus (A) and L. iners (B) are shown in grey and have larger genetic distances compared to the donor-recipient pair. To the right: Overview of microbiome composition in the two participants who converted their microbiome with donor strain engraftment. Sampling date (x-axis) and relative taxa abundance of the taxa (y-axis).

### Adverse events

There were no reports of serious adverse events (SAEs), and none of the adverse events, which was 15 (40.5 %) in the VMT group and 5 (41%) in the placebo group resulted in premature termination of study participation. One participant became pregnant during the study period and subsequently dropped out. The pregnancy was unrelated to the treatment. Another participant was diagnosed with HPV which had not been detected at baseline. This HPV strain was not found in the VMT used in the recipient (see supplementary table 4).

### Flow cytometry analysis

Flow cytometry analysis was conducted to evaluate immune cell populations in both peripheral and menstrual blood in relation to VMT and microbiome conversion. The cell types analyzed included CD56+ natural killer (NK) cells, with subsets CD56^bright^CD16^dim^ and CD56^dim^CD16^bright^, along with their subtypes NKG2A, NKG2D, and NKp46. Additionally, T-cell populations were assessed, including CD8^+^ and CD4^+^ T-cells, regulatory T-cells (Tregs), and T-helper (TH) subsets: TH1, TH2, TH17, TH22. Despite this comprehensive analysis, no significant differences in immune cell populations were observed, in relation to the VMT or dysbiosis resolution – neither in peripheral blood nor in menstrual blood. Results of the flow cytometry analyses are shown in supplementary figure 1a-g and supplementary figure 2a-g.

### Antiseptic pretreatment sub-study

We reasoned that a decrease in the load of vaginal dysbiosis associated bacteria may facilitate the ability of the donor microbiome to engraft and therefore decided to evaluate the effect of antiseptic pretreatment prior to an additional VMT in an open-label extension in refractive participants. We decided to use the antiseptic chlorhexidine, as previous studies have shown its efficacy in decreasing bacterial load (21,22). 27 recipients were refractory to primary treatment (both VMT and placebo) and met the eligibility criteria for and accepted participation in the open-label extension of antiseptic pretreatment. Ten women received pretreatment with antiseptic (Chlorhexidine-Cetrimide 0.2%-0.1%) followed by VMT. Five of ten women (50%) displayed successful conversion of their vaginal microbiome towards a *Lactobacillus-*dominated state at two weeks after treatment. Ten women received a pretreatment with saline water followed by VMT, and seven women received a treatment with antiseptic without VMT. None of these 17 women showed a conversion towards a *Lactobacillus-*dominated microbiome two weeks after treatment. Therefore, antiseptic pretreatment followed by VMT was shown to have a significantly higher conversion rate than saline pretreatment followed by VMT with an odds ratio (OR) of 21 (95% CI 1.83;2956, Bonferroni adj. p-value = 0.033) (figure 5A). SNV analysis of the five women with successful conversion towards a *Lactobacillus-* dominated microbiome showed that at least three of five women showed donor strain engraftment (figure 5B). One did not have enough reads for SNV analysis and one did not show donor engraftment, but conversion.

**Figure 5:**
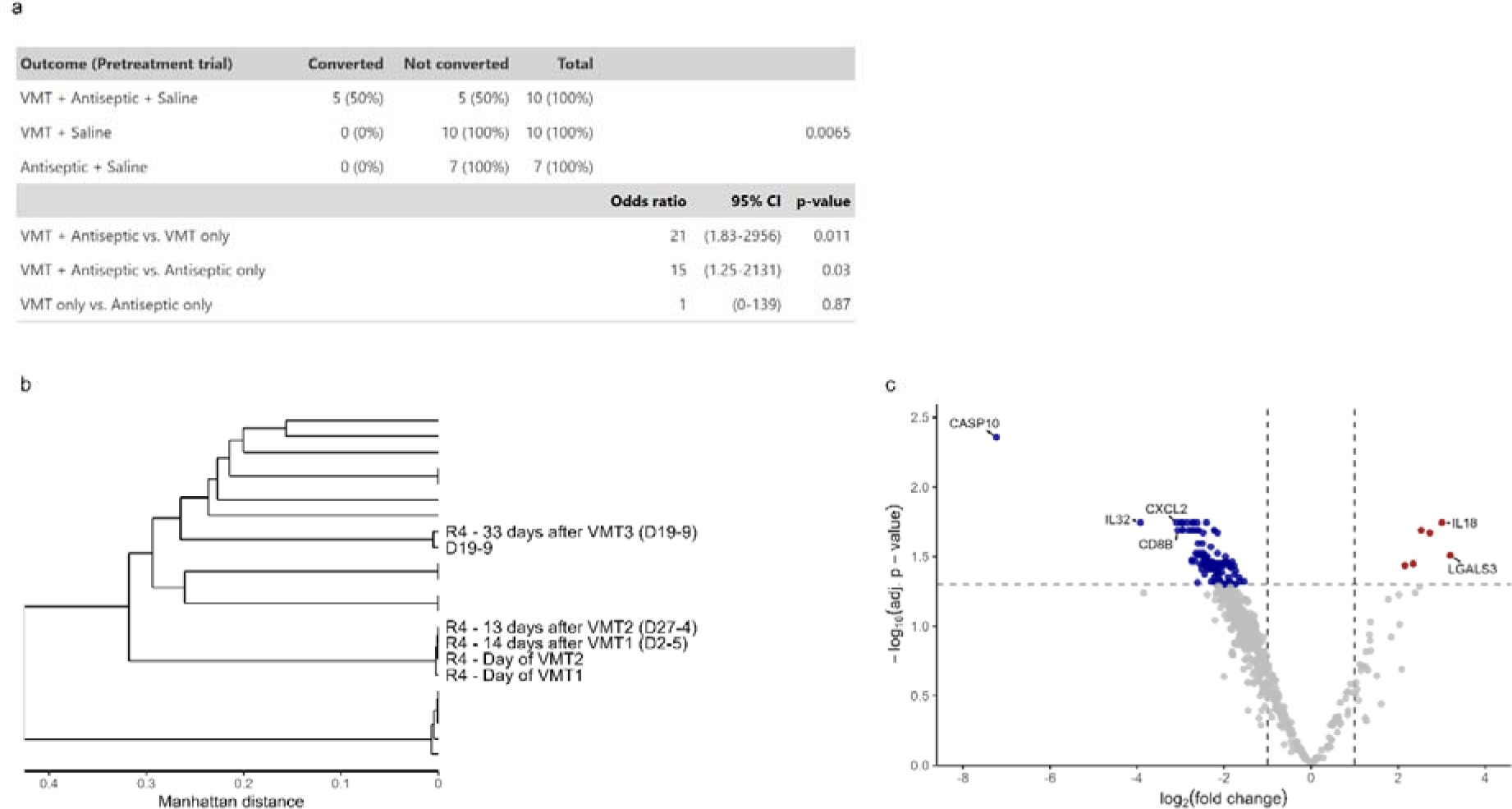
Results from open-label antiseptic pretreatment study A: Odds ratios are for the for comparison of antiseptic treatment + VMT compared to either saline or antiseptic. B: Dendrogram of Manhattan distances between Single Nucleotide Variant (SNV) profiles generated from metagenomic sequencing of CVS samples. The colored text indicates samples from the donor-recipient pair with low genetic distance indicating successful donor engraftment. C: Effects estimates of differentially expressed genes in CVS between converted (n=4) vs. non-converted (n=5) participants in the antiseptic + VMT group after treatment adjusted for pre-treatment expression. Up and down-regulated genes are in red and blue respectively

### KEGG functional analysis of converters and engrafters in both the RCT and antiseptic pre-treatment study

When looking into features, that differed between engrafters and converters, a higher abundance of *L. crispatus* was observed post VMT in recipients with donor engraftment compared to recipients converting their microbiome without donor engraftment indicating a stronger shift towards *lactobacillus* dominance (engrafted: 71-100%, mean 90.1%; not engrafted: 0-100%, mean 32.3%; p = 0.09 after multiple testing adjustment) (supplementary figure 3). Furthermore, functional KEGG analysis of pre-conversion samples show that three pentose-phosphate pathways (PPP) (PENTOSE-P-PWY, NONOXIPENT-PWY, PWY-8178) and one glycolysis pathway (PWY-5484) in energy metabolism were less abundant in the microbiome of the group with donor strain engraftment than in the group who just converted (supplementary figure 3). These pathways were not associated to any particular bacterial species (23).

### Transcriptomics

Using NanoString transcriptomics on cervico vaginal secretion (CVS) in the antiseptic extension study, we identified 130 out of 649 genes that were differentially expressed between the group with successful conversion of the microbiome towards eubiosis after antiseptic + VMT (n=4) when compared to the group that did not convert (n=5) (figure 5C). The group with successful conversion showed significant downregulation in the expression of CASP-10, IL-32, CXCL2, and CD8b. These findings indicate alterations in the gene expression profile post-VMT, with decreased expression levels in genes associated with apoptotic processes (CASP-10), inflammatory response (IL-32 and CXCL2), and cytotoxic T cell activity (CD8b). In addition to the downregulation of certain genes post-VMT, the analysis also revealed significant upregulation in other key genes. Notably, there was an increase in the expression levels of IL-18, a cytokine involved in immune response modulation, and LGALS3, which plays a role in cell adhesion and modulation of cell-cell interactions. Further, using principal component analysis (PCA) on the ratio between pre- and post-VMT expression, we found a distinct clustering of the two groups at PC1 and PC2 (Supplementary figure 4). However, we only observed this effect in three out of four individuals who converted their microbiome (Supplementary figure 5). Based on these analyses, we see distinct changes on a molecular level in response to conversion from dysbiosis to eubiosis after treatment with local antiseptic followed by VMT.

### Discussion

Building upon the pioneering landmark study on the concept of VMT (19), our study, alongside the accompanying paper by Bosma and Mortensen et al., represents the first RCT to evaluate the efficacy of VMT in women with both symptomatic and asymptomatic vaginal dysbiosis in up to three menstrual cycles without prior antibiotic treatment. We could not identify a significant effect of a VMT when given as a single dose per menstrual cycle and without any form of pretreatment. There are, however, two aspects of note: a) the donor strain engraftment in two women who shifted to *Lactobacillus*-dominance was persistent up to the last follow up after 199 days. b) in an open-label extension investigating the effect of antiseptic pretreatment, we showed a significantly higher conversion rate in women treated with the antiseptic + VMT than with antiseptic or VMT alone. Notably, this was in participants who had failed to respond to the treatment in the initial part of the study. Moreover, most women who converted after antiseptic + VMT also showed donor strain engraftment. These first results underscore the potential of VMT to facilitate the engraftment of a *Lactobacillus*-dominated microbiome and thereby resolve vaginal dysbiosis in some recipients. The persistent donor strain engraftment observed supports the conclusion that these were genuine treatment-induced changes rather than spontaneous conversions. In a study previously published by our group, we reported sustained donor strain engraftment for 430 days after VMT without pretreatment in a woman with vaginal dysbiosis and a history of recurrent late pregnancy losses (20). In that study, we performed donor selection based on the ability of the bacteria in donor CVS to inhibit the growth of dysbiosis- associated bacteria in the patient using a plate diffusion assay. This was not performed in this study but could potentially have increased success rates.

The parallel performed trial of Bosma and Mortensen et al. in a recipient population with asymptomatic vaginal dysbiosis found VMT to be more successful than demonstrated here. Of note, in that study the recipients received a VMT on three consecutive days and the definition of when a VMT was successful was defined as the change in overall vaginal lactobacilli (combination of *L. crispatus, L. iners, L. gasseri, and L. jensenii*) by relative abundance and not with a predefined 70% relative abundance of vaginal lactobacilli (combination of *L. crispatus, L. iners, L. gasseri, and L. jensenii*) and a combined proportion of *Gardnerella spp., F. vaginae, and Prevotella spp*. of <10%, as in this study.

The observed changes in gene expression after dysbiosis resolution, including the upregulation of IL-18 and LGALS3 and the downregulation of CASP-10, IL-32, CXCL2, and CD8b, provide insights into the immunological impact of VMT. The upregulation of IL-18 and LGALS3 suggests enhanced immune surveillance and integration of the new microbiota (24,25), while the downregulation of CASP-10, IL-32, CXCL2, and CD8b indicates a shift towards reduced inflammation and cytotoxicity (26–28). These findings highlight VMT’s potential as a modulator of local immune responses, fostering a stable vaginal microbiome, but should be interpreted with caution due to the small sample size.

In contrast to the gene expression changes observed post-VMT, our flow cytometric analysis of both peripheral and menstrual blood revealed no significant differences in the systemic and local immune cell populations. This absence of detectable changes on a cellular level could suggest that the immunological impact of VMT, at least in the context of circulating immune cells, might be more subtle or localized. These findings although based on a limited number of individuals highlight the complex immune responses to VMT and the need for multifaceted research. Further studies on localized tissue responses and longitudinal immune monitoring could deepen our understanding of immune dynamics following VMT.

Our study aimed to identify patterns or characteristics of recipients exhibiting donor strain engraftment and to determine if specific donor VMTs were more effective. Taxonomically, recipients with donor engraftment showed significantly higher levels of *L. crispatus* post-VMT treatment compared to those in the conversion group, suggesting a more substantial shift towards eubiosis. Additionally, functional analysis indicated that recipients with confirmed donor strain engraftment displayed distinct differences in genes associated with energy metabolism, particularly the pentose phosphate pathway (PPP) and glycolysis. While direct research linking these pathways to vaginal health is limited, PPP is crucial for NADPH production, protecting vaginal epithelial cells from oxidative damage, and glycolysis supports the energy needs of these cells, maintaining an optimal environment for beneficial bacteria, primarily lactobacilli (29,30). Since these pathways were more abundant in the subjects that converted without engraftment (in other words, without direct support from the VMT), it could be speculated that they had a healthier baseline state, allowing spontaneous conversion. In contrast, the recipients with donor strain engraftment had a more dysbiotic state at baseline but converted to an even more eubiotic state with a higher *L. crispatus* abundance.

Interesting, when performing VMTs in the antiseptic pretreatment study, we found that some VMTs that did not lead to resolution in the RCT proved to be successful after pretreatment. Overall, we did not observe a clear ‘super donor’ effect, but engraftment seems to be determined by interactions between donor and recipient microbiomes. Future studies including a higher number of subjects are required to reveal the mechanisms determining engraftment of donor microbiomes.

We postulate that the high conversion rate after antiseptic pretreatment is due to a combination of bacterial load reduction and removal or disruption of the vaginal biofilm, which especially *G. vaginalis* is known to produce (31,32). Bacteria within biofilms resist elimination by the immune system and are not completely eradicated by antibiotics, potentially resulting in persistence or recurrence (33–36). It is plausible that the antiseptic Chlorhexidine-Cetrimide weakened the biofilm in participants. This process likely enabled the establishment of a healthy vaginal environment through VMT.

In contrast to a treatment with a single isolated lactobacillus strain, the VMT contains the whole community of multiple strains of lactobacilli, bacteriophages, proteins, cytokines, lipids, antimicrobial peptides that before transplantation formed a stable colonization in the donor. Transplantation of the whole community possibly seems to be key to a lasting engraftment as shown in patients with donor strain engraftment. Among other things, these factors include already resident vaginal bacteria, glycogen and lactic acid (37–39).

While this study marks a significant step forward in exploring VMT as a treatment for vaginal dysbiosis, it also highlights areas for optimization of efficacy. The lack of significant effects without pretreatment with a single dose in our recipient population suggests that factors such as dosing, and donor selection play a crucial role in the efficacy of VMT as a stand-alone treatment. Future studies should consider these variables more closely, potentially also exploring personalized approaches to donor matching when used without antibiotics or antiseptic pretreatment. These refinements could be key in maximizing the therapeutic potential of VMT, making it a more effective and reliable treatment option for vaginal dysbiosis. Replacing dysbiosis-associated bacteria with a *Lactobacillus*-dominated vaginal community and resetting the immunological tone of the reproductive tract could significantly influence a broad spectrum of reproductive and women’s health conditions.

### Strengths and limitations

The clinical trial being conducted in Denmark and reflecting the ethnic makeup of this population limits the generalizability of the findings to other populations, where the frequency of BV has previously been reported to be higher than observed in this study (7,40). Although being the largest VMT study we have limited statistical power in flow cytometry and antiseptic pretreatment sub-studies. Additionally, the post-hoc analysis comparing engrafters versus non-engrafters and their characteristics would require a larger sample size to draw robust conclusions. Overall, the studies presented here highlight crucial fundamental insights into the benefits of VMT and establish essential methodological foundations to support future large-scale intervention studies for various conditions and diseases affecting the female reproductive tract.

## Methods

### Study design

This double-blinded randomized controlled-trial of VMT in women with vaginal dysbiosis, was conducted at Copenhagen University Hospital, Hvidovre, Denmark between June 2021-March 2023.

#### Primary outcome

1. Conversion to a *Lactobacillus*-dominated (eubiotic) microbiome following VMT treatment, given up to three times without antibiotic pretreatment [Time Frame: up to six months from transplantation]

#### Secondary outcomes

1. Conversion to a *Lactobacillus*-dominated (eubiotic) microbiome following one VMT treatment [Time Frame: two weeks from transplantation]
2. Identification of donor microbiome in recipients
3. T- and NK-Cell changes in peripheral and menstrual blood after VMT compared to baseline using flow cytometry
4. In treatment-refractory participants, conversion rate two weeks after antiseptic pretreatment followed by an additional VMT when an antiseptic vaginal wash is performed immediately before the VMT.

### Participant recruitment

Women were recruited through social media, internally at the study site at Copenhagen University Hospital, Hvidovre, and via posters at different educational institutions in the Copenhagen area.

The overall inclusion criteria for study participation were women between the age of 18-40 years who considered themselves healthy, were premenopausal and not pregnant (see complete list of in- and exclusion criteria in supplementary table 3). Participants were further screened and stratified into donors and recipients. A pre-screening questionnaire was developed, in which participating women answered detailed questions such as personal anamnesis, family medical history, life-style factors, sexual history and behavior, contraceptive, and vaginal product usage.

Upon agreeing to participate, participants were asked to come to the study site on their cycle day 10±2 (or any day in case of amenorrhea due to hormonal contraceptives), where they received further information about the study in person, signed the informed consent form and self-collected a vaginal swab sample.

#### Vaginal microbiome collection and stratification for donors and recipients

Participants were instructed to sample 5-6 cm into the vagina and to ensure that the swab would not touch other body parts or surfaces. For clinical sampling, Zymo Collection Swab, 80XX (Zymo research, California US) swabs were used and stored in COPAN (Brescia, Italy) containers. See supplementary materials and methods for details on vaginal microbiome analysis.

After this first screening and based on the sequencing analysis of the vaginal microbiome participants were divided into two groups:

- Potential donors: a vaginal microbiome consisting of >80% relative abundance of vaginal *Lactobacillus* spp. (*L. crispatus, L. jensenii, L. gasseri, L. iners*) and <5% relative abundance of the following vaginal dysbiosis-associated bacteria (*Gardnerella* spp*., F. vaginae and Prevotella* spp.),
- Potential recipients: <10% total relative abundance of *Lactobacillus* spp., and >20% relative abundance of *Gardnerella* spp*., F. vaginae* and *Prevotella* spp.

Participants were provided a window of 40 days to attend the second scheduled visit according to the study protocol. During this visit, eligible donors and recipients underwent a comprehensive medical and gynecological assessment. The exam included vaginal inspection, as well as a vaginal ultrasound scan, a general health check including auscultation of heart and lungs, body weight, heart rate and blood pressure measurements. Standard diagnostic blood and vaginal swab tests were conducted to evaluate the presence of various pathogens and conditions, including human immunodeficiency virus (HIV), Hepatitis A/B/C, cytomegalovirus (CMV), and herpes simplex virus (HSV) 1/2 (for a comprehensive list, see supplementary figure 6a & 6b and supplementary tables 1 & 2).

### Intervention procedure in RCT

Recipients needed to be willing to receive up to three VMTs dosed as single dosage once per menstrual cycle, and to refrain from using intravaginal products and avoid activities like pool and bathtub usage. Additionally, eligible recipients were also asked to refrain from vaginal and anal intercourse during the study period, unless using condoms.

Exclusion criteria for recipients were documented disorders such as HIV, or if testing positive for HIV, chlamydia*, Mycoplasma genitalium*, trichomonas or having a positive pregnancy test based on urine human chorionic gonadotropin (HCG) at the second screening visit. See supplementary table 3 for further exclusion criteria.

### Adverse events (AE)

AE was defined as worsening of vaginal symptoms reported by the participants throughout the 6 months study period, serological (HIV, Hepatitis A/B/C, CMV, CRP), sexually transmitted infections (*Mycoplasma*, chlamydia, trichomonas, *Neisseria gonorrhea*, HSV 1/2) and pregnancy. Aliquots of each donor transplant were stored for testing in case of the AE potentially being related to the donor transplant.

### Randomization

At the second visit, participants were randomized to receive the active treatment or placebo in a ratio of 3:1. The randomization was done by the collaborating partner Freya Biosciences ApS (Copenhagen, Denmark), using a computerized randomization program (Sealed Envelope^TM^ Ltd, London, England). We used block randomization with each block consisting of n=4 participants to ensure balanced allocation across treatment groups.

### Procedure

For comprehensive details on donor participant recruitment, as well as sample collection, processing, analysis, and the characterization methods used for both donor and recipient samples see supplementary materials and methods as well as supplementary table 3.

#### Study visits

Women who met the criteria for recipients at the initial screening and successfully met the additional inclusion and exclusion criteria at the second visit were considered suitable as recipients. After the second screening visit, a personalized calendar of future appointments was established based on the participants’ menstrual cycle.

The women were seen every 14 days. On menstrual cycle day (CD) 10 ±2 days, they received the VMT or placebo (see supplemental materials and methods). On CD 24±2 in each menstrual cycle they came into the clinic for a general health check-up, self-performed vaginal swab test, urine test and CVS samples which were collected with a single-use menstrual disc. If recipients had amenorrhea due to hormonal contraceptives, a cycle of 28 days was calculated, and visits planned accordingly.

To facilitate continuous monitoring, an online self-reported questionnaire was implemented. This questionnaire was designed to be completed by recipients one day before each visit. It covered aspects such as recent menstruation, vaginal discharge, sexual activity, condom use, and any new medications or relevant information such as potential side-effects from the treatment.

#### Definition of dysbiosis resolution

Dysbiosis resolution was defined as a minimum of 70% total relative abundance of *Lactobacillus* spp. and a combined proportion of *Gardnerella* spp*., F. vaginae, and Prevotella* spp. of <10%, by shotgun metagenomic sequencing of a vaginal sample.

In total, recipients came into the clinic for nine visits, which could involve up to three VMTs, depending on the success and timing of dysbiosis resolution. Upon dysbiosis resolution, the next scheduled appointment for VMT was changed to a general check-up visit (see supplementary table 2 and supplementary figure 6b). After the third cycle, and therefore final VMT, recipients returned for a follow-up visit after one and three months. Further information on study visits for recipients is provided in supplementary table 2 and supplementary figure 6b.

### Flow cytometry

The first 24 participants with regular menstruation were asked to participate in the flow cytometry study. The participants collected their menstrual blood for the first 48 hours of menstruation for flow cytometry immune cell analysis of menstrual and peripheral blood. There were two timepoints of collection: The first timepoint before the first VMT and a second timepoint after the follow-up 2 visit for the whole population. Thereby, comparing participants with and without conversion of their microbiome (see supplementary table 2 and supplementary figure 7). For further information on cell isolation and procedure and analysis of isolated lymphocytes, see the supplementary materials and methods flowcytometry section.

### Antiseptic pretreatment

Recipients who did not meet the criteria for successful dysbiosis resolution after three single-dose VMTs over three menstrual cycles were invited at the 3-month follow-up visit to participate in an open-label extension of the RCT. Participants were randomly assigned to one of three treatment groups: 1) pretreatment with Chlorhexidine-Cetrimide 0.2%-0.1%, followed by saline, followed by VMT, 2) pretreatment with saline followed by VMT, 3) pretreatment with Chlorhexidine-Cetrimide 0.2% - 0,1% followed by saline and no VMT.

Each participant was scheduled for two visits: The initial visit was planned for CD 10±2, and consisted of a general health-check and collection of biobank samples (see supplementary table 1b and supplementary figure 8). For a description of procedure in the three treatment groups see supplementary material and methods Procedure in the three treatment groups in the open-label extension section.

On CD 24±2, participants had a general health check and self-collected vaginal swab, urine, and CVS samples. Vaginal swab samples were tested for *Chlamydia trachomatis, N. gonorrhea, Trichomonas vaginalis, M. genitalium, group A, B, C & G Streptococcus, HSV 1/2* and *Candida* spp. Additionally, a gynecological examination and ultrasound scan were performed and potential side effects, if any, were registered. For further information on the antiseptic pretreatment see table 2 and figure 4 in supplementary materials and methods. To identify changes in the immunological response at a transcriptional level, CVS was collected at each visit and analyzed using the NanoString nCounter Human Immunology V2 Panel (see supplementary materials and methods transcriptomic section for details).

### Statistical analysis

Sample size calculation was based on results from the only existing intervention study on VMT, which reported an 80% remission rate of BV (19). A 3:1 randomization was chosen to balance the considerations of randomized design while optimizing the number of VMT treatments, yielding for a more robust and nuanced understanding of its efficacy and risks.

Using these parameters and aiming for 80% statistical power to detect a significant difference (p<0.05), our sample size calculation indicated that a total of 48 participants (36 recipients in the active treatment arm and 12 in the placebo arm) were required.

We estimated that it would be necessary to screen 300-350 women to find 25 donors and 48 recipients, given that 16-20% of Danish women of fertile age have been shown to have vaginal dysbiosis (5).

For demographic data, counts with percentages and means with standard deviations are reported. For primary and secondary outcomes, Fisher’s exact test was used for odds ratios, confidence interval and p- values. We used unadjusted Kaplan-Meier curves and the log rank test to compare survival patterns by VMT and placebo groups. All analysis was conducted in R (v. 4.3.0)

Fisher’s exact test was used to determine the independence between the three groups in the antiseptic pretreatment study. Group-by-group comparisons were conducted by Firth logistic regression for small sample analysis using the logist package in R (v. 1.26.0). P-values were adjusted using Bonferroni correction.

The primary outcome was based on Intention To Treat (ITT). Since all participants followed the treatment group they were assigned at randomization a per protocol analysis was not performed.

See supplementary materials and methods for details on microbiome and flow cytometry statistical data as well as transcriptomic statistical data.

No data monitoring committee was assigned to this study.

## Legal and ethics statements

The Randomized Controlled Trial study protocol was approved by the Danish ethical comittee of the capital region of Denmark (Journal-nr.: H-20068230) on the 16th of March 2021 and the Danish Data Protection Agency (Pactius): P-2020-1080. The Randomized Controlled Trial was reported on ClinicalTrials.gov Identifier: NCT04855006.

Three amendments were added to the study protocol: first, adding rectal swab collection (approved by the Danish ethical Committee of the capital region of Denmark on the 15th of June 2021), second, sub-study using flow cytometry to study immunological cells in peripheral and menstrual blood ( approved by the Danish ethical Committee of the capital region of Denmark on the 6th of December 2021) and third, a sub- study of the impact of antiseptic pretreatment to the participants who did not meet the criteria for successful VMT (approved by the Danish ethical Committee of the capital region of Denmark on the 17th of May 2022).

All research was conducted according to the Declaration of Helsinki and all participants provided written informed consent.

## Other information

### Data availability statement

The datasets generated during and/or analyzed during the current study are available from the corresponding author on reasonable request.

### Funding

The study was partially funded (i.e., analysis costs) by Freya Biosciences Aps, Fruebjergvej, 2100 Copenhagen, Denmark.

### Conflict of interest

Elleke F. Bosma, Brynjulf Mortensen, Kevin DeLong, and Johan E.T. van Hylckama Vlieg are employees of Freya Biosciences, ApS. Johan E.T. van Hylckama Vlieg, Laura M. Ensign, and Anne Bloch Thomsen are co- founders and shareholders of Freya Biosciences, ApS. Laura M. Ensign is a co-inventor on patents and patent applications in the area of microbial transplantation. Laura M. Ensign’s arrangements have been reviewed and approved by the Johns Hopkins University in accordance with its conflict-of-interest policies. All other authors declare that they have no competing interests.

### Contributions

Designed the study:

Henriette Svarre Nielsen, Tine Wrønding, Brynjulf Mortensen, Laura M. Ensign, Johan E.T. van Hylckama Vlieg Kilian Vomstein and Nina La Cour Freiesleben

Participant recruitment, clinical performance and sampling:

Donors:

Elleke F. Bosma, Brynjulf Mortensen, Tine Wrønding, Ann Marie Hellerung, Klara Dortea Wiil, Emilie Vester Engel and Kilian Vomstein

Recipients:

Tine Wrønding, Ann Marie Hellerung, Klara Dortea Wiil, Emilie Vester Engel and Kilian VomsteinWrote first version of the manuscript:

Tine Wrønding, Kilian Vomstein and Henriette Svarre Nielsen.

Revising the manuscript critically for important intellectual content:

Tine Wrønding, Kilian Vomstein, Henriette Svarre Nielsen, Elleke F. Bosma, Brynjulf Mortensen, Henrik Westh, Julie Elm Heintz, Sara Mollerup, Laura M. Ensign, Kevin DeLong, Johan van Hylckama Vlieg, Anne Bloch Thomsen, Andreas Munk Petersen, Ann Marie Hellerung, Klara Dortea Wiil, Emilie Vester Engel, Sofie Ingdam Halkjær, Tanja Schlaikjær Hartwig, Agnete Troen Lundgaard, David Westergaard, Nina La Cour Freiesleben, Luisa W. Hugerth

Performed the vaginal microbiome analysis and bioinformatics:

Julie Elm Heintz, Sarah Mollerup, Agnete Troen Lundgaard, David Westergaard, Kevin DeLong, Henrik Westh, Andreas Munk Petersen and Luisa W. Hugerth

All authors made substantial contributions to the study set-up, data interpretation and manuscript revision. All authors approved the final manuscript before publication.

## Notes

### Clinical Trial

NCT04855006

### Author Declarations

The Randomized Controlled Trial study protocol was approved by the Danish ethical comittee of the capital region of Denmark (Journal-nr.: H-20068230) on the 16th of March 2021 and the Danish Data Protection Agency (Pactius): P-2020-1080. The Randomized Controlled Trial was reported on ClinicalTrials.gov Identifier: NCT04855006. Three amendments were added to the study protocol: first, adding rectal swab collection (approved by the Danish ethical Committee of the capital region of Denmark on the 15th of June 2021), second, sub-study using flow cytometry to study immunological cells in peripheral and menstrual blood (approved by the Danish ethical Committee of the capital region of Denmark on the 6th of December 2021) and third, a sub-study of the impact of antiseptic pretreatment to the participants who did not meet the criteria for successful VMT (approved by the Danish ethical Committee of the capital region of Denmark on the 17th of May 2022). All research was conducted according to the Declaration of Helsinki and all participants provided written informed consent.

