## Supplementary materials and methods for "Vaginal Microbiota Transplantation (VMT) for treatment of vaginal dysbiosis without the use of antibiotics – A Double-Blinded Randomized Controlled Trial in healthy women with vaginal dysbiosis"

Donors

In addition to the microbiome and clinical characterization, participation as donor required the absence of vaginal symptoms indicative of dysbiosis, such as malodorous discharge of yellow/greenish color or vaginal itchiness. Donors also needed to be willing to donate 10 cervicovaginal secretion (CVS) samples over a period of up to 40 days, and to refrain from using intravaginal products and avoid activities like pool and bathtub usage. Additionally, donors were asked to refrain from vaginal and anal penetration unless using condoms during the entire donation period.

Exclusion criteria for donor participation included a history of vaginal health issues such as BV, trichomonas, syphilis, human papillomavirus (HPV)-related conditions (cervical cell changes, genital warts, herpes), recurrent cystitis, and *Mycoplasma genitalium*. Severe infections, such as HIV and hepatitis also led to exclusion. Recent (within the last year) cases of gonorrhea or chlamydia, current pregnancy or breastfeeding, use of specific medications (antibiotics, hormonal treatments other than hormonal contraceptives), high-risk sexual behaviors for STIs (defined as many sexual partners and unprotected intercourse), and recent (last 3 months) IUD removal or cervical laser treatment were additional exclusion criteria. Detailed information on all exclusion criteria can be found in supplementary table 3. Further information on and illustration of study visits for donors are shown in supplementary table 1 and supplementary figure 1. These criteria are similar to the donor program describes in our accompanying paper by Bosma & Mortensen et al. except donors were not allowed to have any intercourse at all and could also not swim in the ocean during the donation period in the accompanying study.

**Donor CVS processing and quality control:**

Donor CVS sampling and processing was performed as described in the accompanying paper by Bosma & Mortensen et al.

**Intervention procedure in RCT**

Recipients needed to be willing to receive up to three VMTs dosed as single dosage once per menstrual cycle, and to refrain from using intravaginal products and avoid activities like pool and bathtub usage. Additionally, eligible recipients were also asked to refrain from vaginal and anal intercourse during the study period, unless using condoms.

Exclusion criteria for recipients were documented disorders such as HIV, or if testing positive for HIV, *Chlamydia*, *Mycoplasma genitalium*, *Trichomonas* or having a positive pregnancy test based on urine human chorionic gonadotropin (HCG) at the second screening visit. See supplementary table 3 for further exclusion criteria.

Study visits

Women who met the vaginal microbiome criteria for recipients at the initial screening and successfully met the additional inclusion and exclusion criteria at the second were considered suitable as recipients. After the second screening visit, a personalized calendar of future appointments was established based on the participants’ menstrual cycle. Participants were encouraged to communicate if any changes in their cycle would impact their scheduled appointments.

The women were seen every 14 days. On cycle day (CD) 10 ±2 days, they received the VMT or placebo (see supplemental materials and methods). On CD 17±2, they performed a self-swab at home and sent it on the same day with express mail to the study site. On CD 24±2 in each menstrual cycle they came into the clinic for a general health check-up, self-performed vaginal swab test, urine test and CVS samples which were collected with a single-use menstrual disc. Biobank (plasma, urine, CVS, rectal swab) samples were collected during the baseline visit and continued to be taken at all subsequent visits. An overview of samples taken at each study visit is shown in supplementary table 2 for recipients and supplementary figure 1B for recipients. If recipients had amenorrhea due to hormonal contraceptives, a cycle of 28 days was calculated, and visits planned accordingly.

To facilitate continuous monitoring, an online self-reported questionnaire was implemented. This questionnaire was designed to be completed by recipients one day before each visit. It covered aspects such as recent menstruation, vaginal discharge, sexual activity, condom use, and any new medications or relevant information such as potential side-effects from the treatment.

**Intervention procedure:**

Both the VMT product and the placebo product, which consisted of saline water, were stored in a cryovial in a –80°C freezer. The thawing procedure consisted of: 1 hour at room temperature followed by 30 min in a 37°C heating cabin. The VMT product was then drawn up into a 10 mL syringe affixed with a 180 mm Intrauterine insemination catheter (Wallace, 180mm Catheter, Cooper Surgical, CT, USA). The catheter was then introduced into the vagina (to the fornix) without the use of a speculum and the content of the syringe was applied. To ensure a complete emptying of the catheter, an additional 0,5 mL of air was drawn into the syringe and applied as well.

To ensure double blinding, the syringe containing the transplant/placebo was concealed with colored tape by a separate research assistant preparing for the procedure, so neither the participant nor the research assistant performing the VMT could observe the content of the syringe. The research assistant who prepared the samples had no other task in the study, i.e. handling of data or interpretation of data.

**Recipient CVS sample processing and pH measurement**:

CVS samples were obtained through self-collection using a menstrual cup (Softdisc menstrual cup by The Flex Company, also sold under the name ‘Flex disc’) after thorough instructions. The participants wore sterile gloves to insert the menstrual cup longitudinally into the vagina, left it in place for 10 sec, and then twisted the cup while removing it. The patient then placed the CVS-coated menstrual cup into a 50 mL Falcon tube (Corning, US). The 50 mL tube was centrifuged (Eppendorf Centrifuge 5810 R) for 5 min at 190 xg at 4°C to collect the CVS. The menstrual cup was then removed, and the 50 mL tube was re-weighed. 2.5 µL of sterile saline was added per mg sample. The diluted sample was aliquoted into cryovials for biobanking and DNA extraction. An amount of 150 µL of diluted sample was placed in an Eppendorf tube for pH measurement and set aside at room temperature for 15 min. Using a pH-meter (VWR pHenomenal pH 1100L with VWR pH electrode pHenomenal MIC 220) the pH was measured three times, and the mean pH was used for further analyses.

**Vaginal microbiome analysis/extraction:**

1 mL of each vaginal swab sample or 150 µL CVS was used for DNA extraction with MolYsis™ Complete5 DNA extraction kit (Cat# D-321-100; Molzym GmbH & Co. KG, Bremen, Germany). Samples were processed according to manufacturer’s instruction with following alterations: lysis of bacteria in a thermoshaker at 250 rpm for 30 min and 37 °C (β-mercaptoethanol not added), followed by Proteinase K treatment in a heating block at 56 °C for 10 min (samples were shortly vortexed and placed back after 3 min and 7 min).

Eluted DNA was stored at 5 °C for a maximum of 24 h before initiation of library preparation. DNA libraries were prepared using the Nextera XT DNA Sample Preparation Kit (Illumina) according to the manufacturer’s instructions and sequenced on a MiSeq (Illumina Inc., San Diego, USA) using 2 x 150 bp paired-end reads.

**Microbiome bioinformatics:**

The quality of the fastqc reads before and after quality trimming was assessed with FastQC v. 0.11.8 (1). Reads were quality trimmed according to qualified_quality_phred of 20 and minimum read length of 50 using fastp v. 0.20.1 (2). The reads were depleted of human sequences by aligning the trimmed reads to the human genome (hg38, University of California, Santa Cruz) using bowtie2 v. 2.3.4.1 (3) with end-to-end alignment and a maximum fragment length for valid paired-end alignments (-X) of 2000. Microbial profiling of the human DNA depleted reads was performed with Kraken 2 v. 2.1.2 (4) using the PlusPF database available from https://benlangmead.github.io/aws-indexes/k2 which includes RefSeq genomes of archaea, bacteria, viruses, plasmids, human, UniVec Core, protozoa, and fungi. Species and genus level abundances were estimated from the Kraken 2 reports using Bracken v. 2.6.2. The relative abundance of bacterial species and genera detected was calculated relative to the total number of reads classified as bacterial. Species-level annotation was performed with Metaphlan4(5), and gene and pathway functional annotation was performed with Humann3(6). Differential abundance analysis was performed in R using the library Aldex2 (7). The p-value considered is from Welch’s t-test. With the exception of *L. crispatus* abundance post VTM, p-values presented are not corrected for multiple testing.

**Single Nucleotide Variant (SNV)-analysis:**

Samples at all timepoints where the coverage was high enough to generate SNV profiles before and after. *L. crispatus* SNV profiles were generated using MetaSNV under default settings, using a publicly available NCBI L. crispatus genome (NCBI accession number CP118069) to generate the reference database. Reads were first aligned to the database using Bowtie2 and filtered to exclude alignments smaller than 45 base pairs and with less than 97% identity. Filtered reads were then used to generate the SNV profile of each metagenomic sample and Manhattan distances were calculated between profiles.

**Flowcytometry**

Sample Collection Procedures

Peripheral Blood (PB):

18 mL of peripheral blood was collected in Vacuette® EDTA tubes (Greiner Bio-One, Kremsmünster, Austria), on the second day of the menstrual cycle.

Menstrual Blood (MB):

Participants gathered menstrual blood during the initial 48 h of menstruation utilizing a menstrual cup (AllMatters ApS, Copenhagen, Denmark) into two falcon tubes for distinct analysis of each 24-hour segment. These tubes were supplemented with 10 mL RPMI 1640 media containing 10% fetal bovine serum (FBS, Sigma-Aldrich, St. Louis, MO, USA), 0.3% sodium citrate (Merck), pyruvate (1 mM), penicillin (100 U/mL), and streptomycin (100 g/mL) (Thermo Fisher Scientific, Waltham, MA, USA).

Cell Isolation

Peripheral Blood (PB):

PB mononuclear cells (PBMC) were isolated employing Lymphoprep density gradient centrifugation (STEMCELL Technologies, Vancouver, BC, Canada) as per the manufacturer's instructions, using Leucosep tubes (Greiner bio-one).

Menstrual Blood (MB):

Prior to cell isolation, each MB sample underwent assessment for volume, viscosity, coagulation, and potential cell lysis. Mononuclear cells (MMC) were isolated from MB after an initial washing and straining step through a 150 µm cell strainer (pluriStrainer®, pluriSelect Life Science, Leipzig, Germany). This was followed by density gradient centrifugation (STEMCELL Technologies, Vancouver, BC, Canada), with subsequent washing steps as directed by the manufacturer.

Analysis of Isolated Lymphocytes and NK Cell Subsets:

Counting of PB, MB, and EB-cells was conducted in a Neubauer chamber, followed by washing in DPBS-FBS 1%. We utilized 5 million cells per sample (300µl). Cells were blocked using Fc Blocking Reagent (Miltenyi Biotec, Bergisch Gladbach, Germany) and stained with antibody master mixes. These mixes comprised the following antibodies: CD16, CD45, CD3, CD4, CD8, CD64, CD14, CD19, CD57, CD62L, CD335 (NKp46) (BD Biosciences, San Jose, CA, USA), CD127, CD25 (BioLegend, San Diego, CA, USA), CD56, CD314 (NKG2D) (Miltenyi Biotec), for 20 minutes at room temperature. After staining with 7AAD (BD Biosciences), samples were processed through a BD LSRFortessa flow cytometer (BD Biosciences, San Jose, CA, USA). Data acquisition was halted upon reaching 150,000 CD45+ cells. Daily calibration with CST beads (BD Biosciences) was performed to minimize inter-experimental variability. FlowLogic software (Inivai technologies, Mentone, Victoria, Australia) was employed for data analysis (refer to supplementary Figure XX for gating strategy). NK-cell receptor analysis was performed on CD3-CD56dimCD16bright and CD3-CD56brightCD16dim NK cell populations.

**Procedure in the three treatment groups in the open-label extension:**

For participants assigned to the first group (pretreatment with Chlorhexidine-Cetrimide 0.2% - 0.1% followed by saline, followed by VMT), the procedure was as follows: The vaginal area was dried using dry gauze, followed by washing of the vagina and fornix with Chlorhexidine-cetrimide and then saline. After another drying step, the active transplant was applied following the VMT procedure. Participants in the second group (saline followed by VMT) received vaginal and fornix washing with saline only, followed by the VMT. The individuals randomized to the third group (pretreatment with Chlorhexidine-Cetrimide 0.2% - 0.1% followed by saline) underwent washing only.

**Transcriptomics– NanoString**:

Differential expression (DE) analysis

Gene expression of CVS was quantified using the Nanostring nCounter Human Immunology V2 Panel with additional 55 custom genes. Differential expression (DE) analysis was conducted using the DESeq2 package (v. 1.40.2) with the design ~status2 week follow-up + treatment + ID using pre- and post-treatment samples from individuals receiving VMT and antiseptic treatment + saline. In this design matrix, status2 week follow-up signifies whether an individual has successfully converted to a *Lactobacillus*-dominated vaginal microbiome (> X % lacto X,Y,Z) two weeks after transplantation, treatment indicates whether the sample was taken pre or post treatment, and ID signifies the individual. The data was normalized to housekeeping genes using the estimateSizeFactors function before running DESeq2 after log2-transformation. DE was based on comparison between samples at post-treatment and p-values were adjusted for multiple testing using False Discovery Rate (FDR) set at alpha = 0.05. All other DESeq2 parameters were set to the default settings. DE genes (DEGs) with an FDR-adjust p-value below 0.05 and a log2(fold change) above 1 or below -1 was defined as significantly up- or down-regulated, respectively.

Gene enrichment analysis

Gene enrichment analysis DEGs was performed using WebGestaltR (v. 0.4.6) using over-representation analysis (ORA). DEGs s were compared to all measured genes using ensembl gene IDs. Mapping of RefSeq mrna IDs as provided by Nanostring to ensembl gene IDs was done using biomaRt (v. 2.56.1). We observed considerable changes in bacterial load after successful conversion in the antiseptic pretreatment study which may influence the normalization to housekeeping genes. However, validation of these data shows a limited effect of bacterial load on gene to housekeeping gene-ratio (data not shown).

Clustering analysis

For the clustering analysis, gene expression data was normalized using DESeq2 with the design ~1, normalized to positive and negative control genes as stated above. For post-treatment/pre-treatment-ratio data, the pre-treatment log2-transformed expression was subtracted from the post-treatment log2-transformed expression. To create heatmaps for the three time points, as well as post-treatment/pre-treatment-ratios, hierarchical clustering was performed on Euclidean-distances from scaled data using the hclust (method = “complete”) and dist functions from the stats package (v. 4.3.0). Dendrograms was created using the as.dendrogram function from the stats package (v. 4.3.0) and extracted using the dendro_data frunction from the ggdendro package (v. 0.1.23). Heatmaps were created using the ggplot2 package (v. 3.4.3). The percentage of lactobacillus was calculated at the sum of fractions of Lactobacillus crispatus, Lactobacillus gasseri, Lactobacillus iners, and Lactobacillus jensenii. Principal component analysis (PCA) was performed using the prcomp function from the stats package (v. 4.3.0) with scaling and centering and plotted using the ggplot2 package (v. 3.4.3).
