## Supplementary figures for "Vaginal Microbiota Transplantation (VMT) for treatment of vaginal dysbiosis without the use of antibiotics – A Double-Blinded Randomized Controlled Trial in healthy women with vaginal dysbiosis"

Figure 1a-g: Main lymphocyte subsets in peripheral and menstrual blood before and after VMT.

a) CD4+ T-cells, b) CD8+ T-cells, c) CD19+ B-cells, d) CD56+ NK-cells, e) CD3+CD56+ NK cells (all shown as % of CD45+ cells), f) CD56brightCD16dim NK-Cells, g) CD56dimCD16bright NK cells (both displayed as % of CD56+ cells). Connected dot plots showing the individual data points of each participant. To the left and right boxplots indicating the median, the quartiles, the range of values in each respective group.

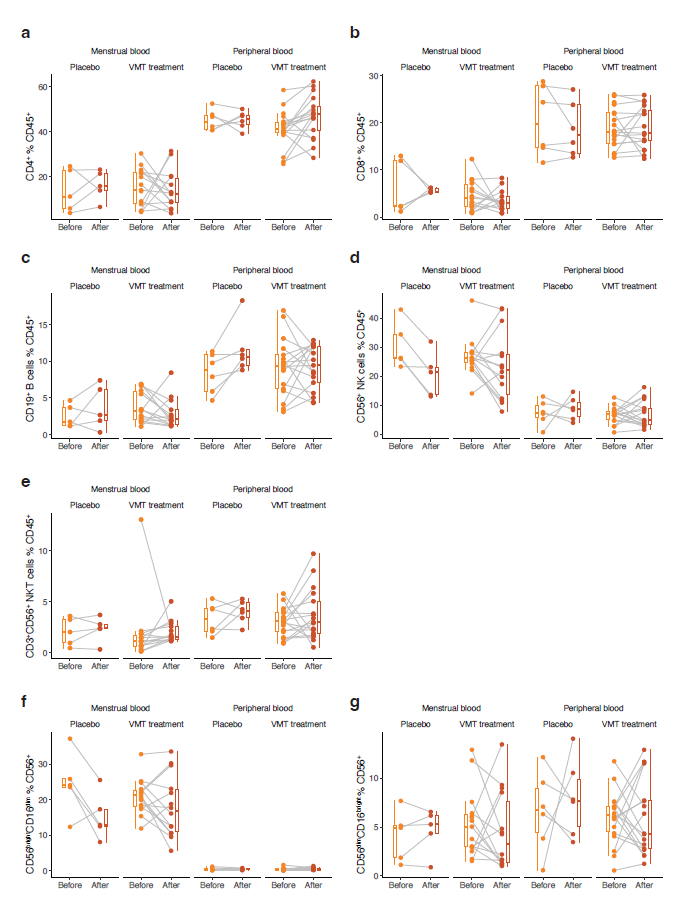

Figure 2a-f: T-cell subsets in peripheral and menstrual blood before and after VMT.

a) Th1-cells, b) Th2-cells, c) Th9-cells, d) Th17-cells, e) Th22-cells f) Treg CD25+CD127low/neg (all CD4+, shown as % of CD45+ cells). Connected dot plots showing the individual data points of each participant. To the left and right boxplots indicating the median, the quartiles, the range of values in each respective group.

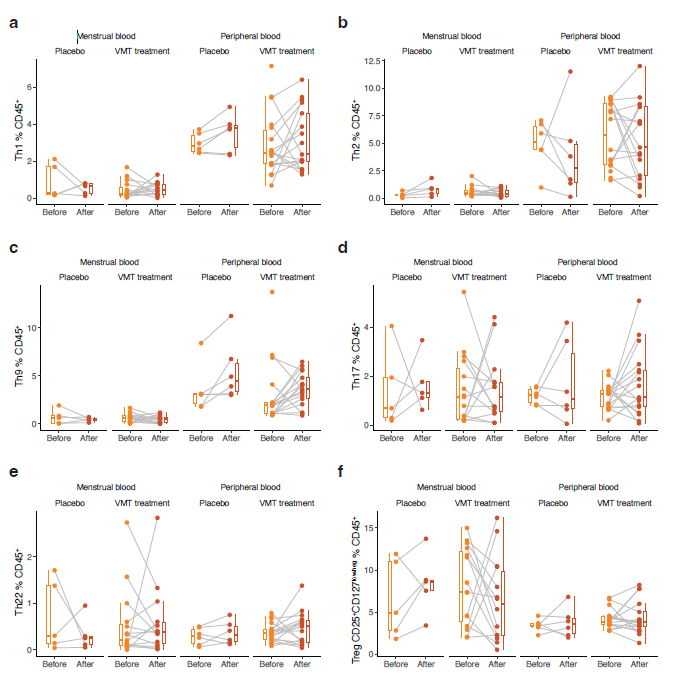

Figure 3

Effect size of change between non-engrafting converters (left) and engrafters (right), based on Aldex2. (a) Species abundance at the first sample after conversion. (b) Pathway abundances before conversion. Dark blue indicates Welch's test p-value <0.05 and light blue indicates p-value 0.05-0.1. THRESYN-PWY: L-threonine biosynthesis. PENTOSE-P-PWY: pentose phosphate pathway. NONOXIPENT-PWY: pentose phosphate pathway (non-oxidative branch) I. PWY-8178 pentose phosphate pathway (non-oxidative branch) II. PWY0-1296: purine ribonucleosides degradation. PHOSLIPSYN-PWY: super pathway of phospholipid biosynthesis I (bacteria). PWY-5121: superpathway of geranylgeranyl diphosphate biosynthesis II (via MEP).

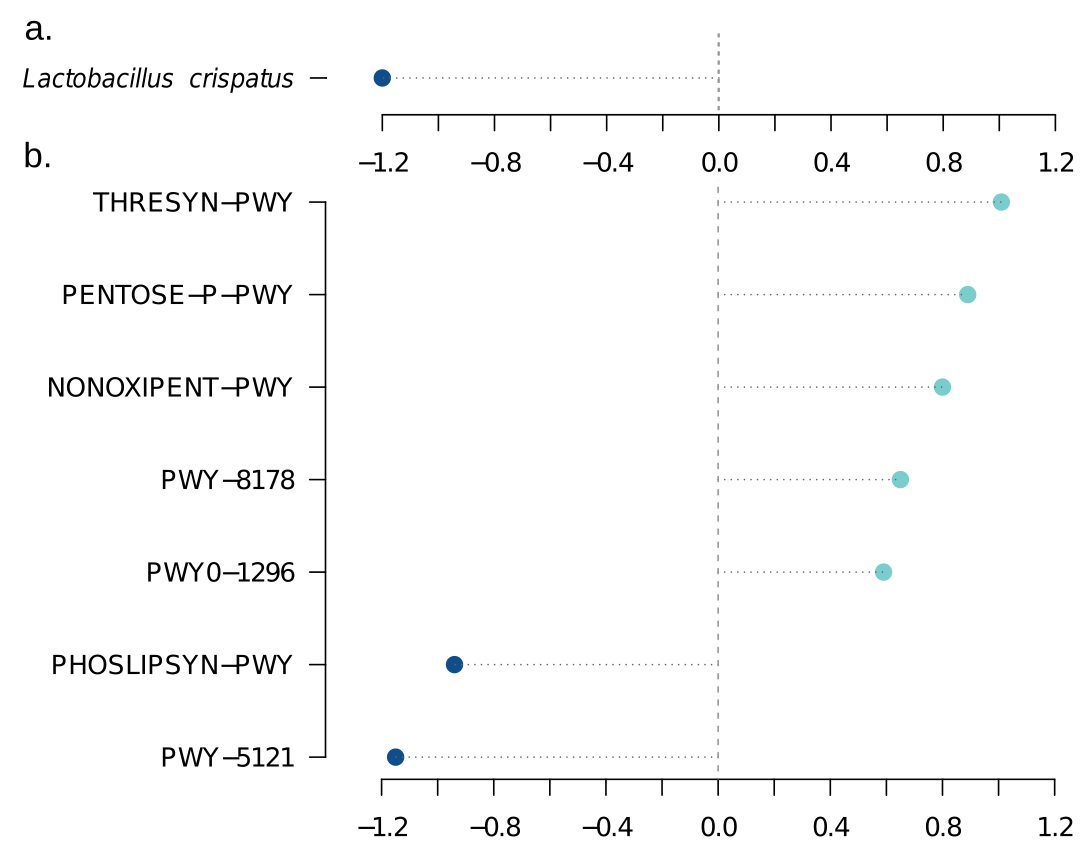

Figure 4

Principal component analysis (PCA) of change in gene expression from pre-treatment to post-treatment. Red and blue dots indicate treatment outcomes in the antiseptic + VMT group, while grey indicates samples from individuals in the VMT-only and antiseptic-only groups.

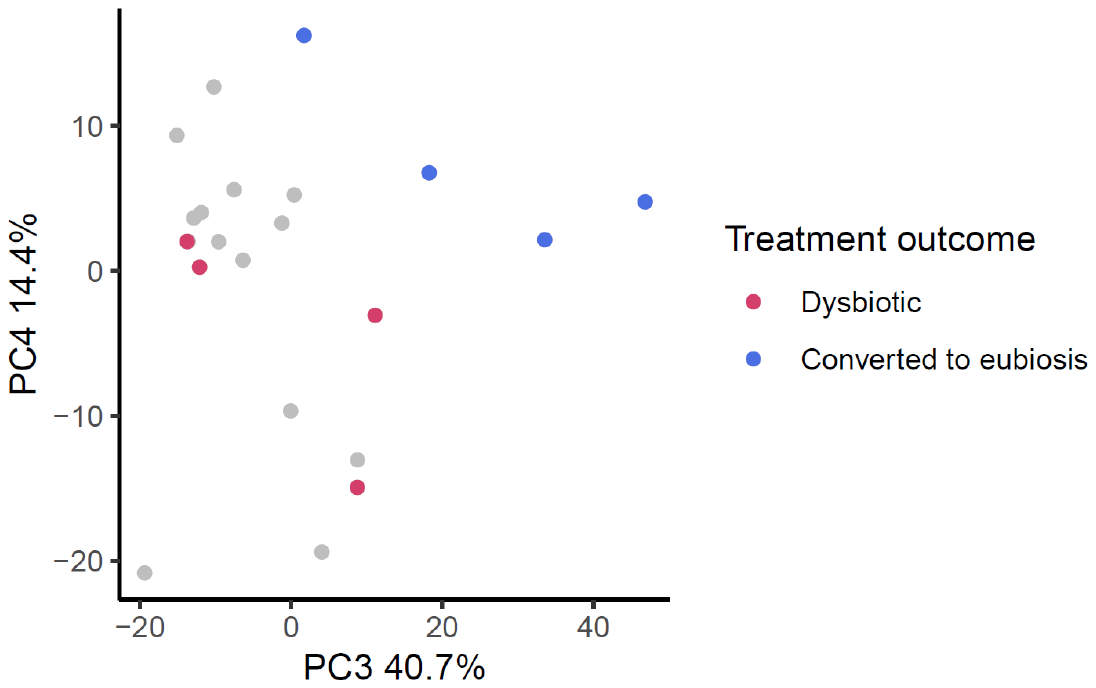

Figure 5

Heatmap of gene expression levels clustered by genes (x-axis) and participants (y-axis). Expression levels are Z-score normalized. Treatment groups and the treatment outcomes for the antiseptic + VMT group are shown on the left.

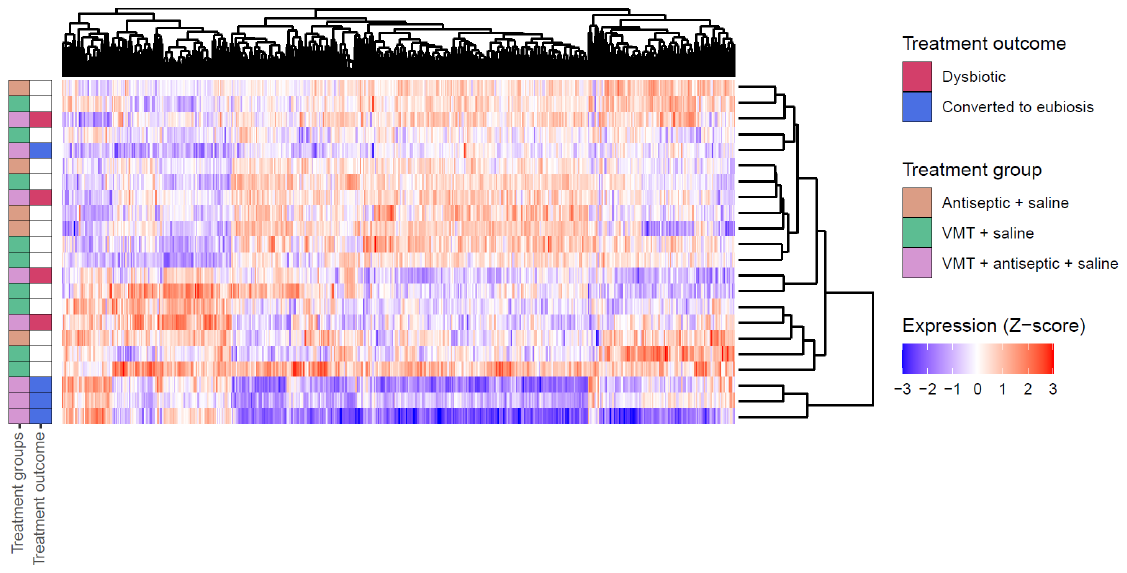

Figure 6a

Study overview of participants and timeline of sample collections for women verified as donors in RCT study. Created with BioRender.com. CD, cycle day.

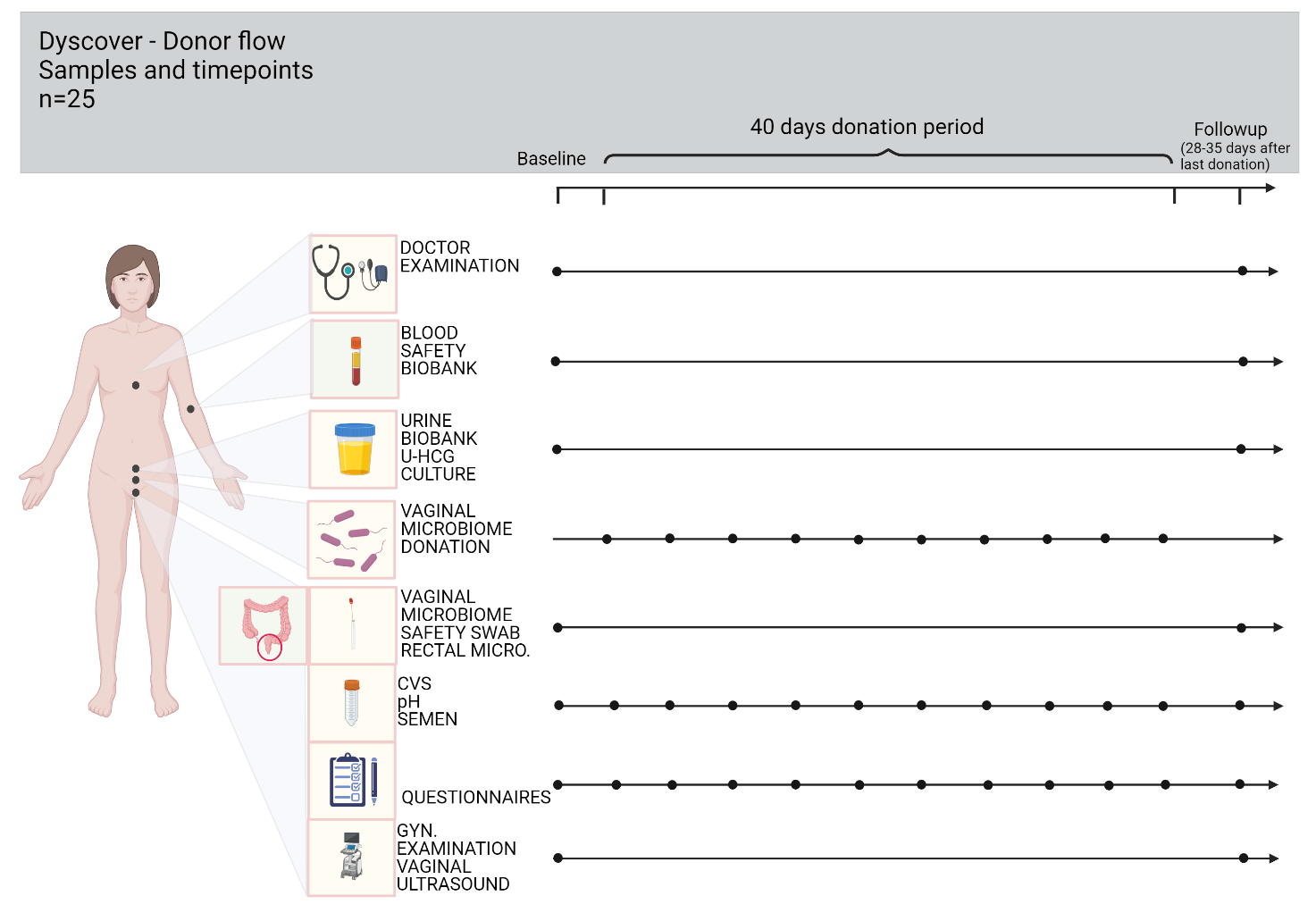

Figure 6b.

Study overview of participants and timeline of sample collections for women verified as recipients in RCT study. Created with BioRender.com. CD, cycle day.

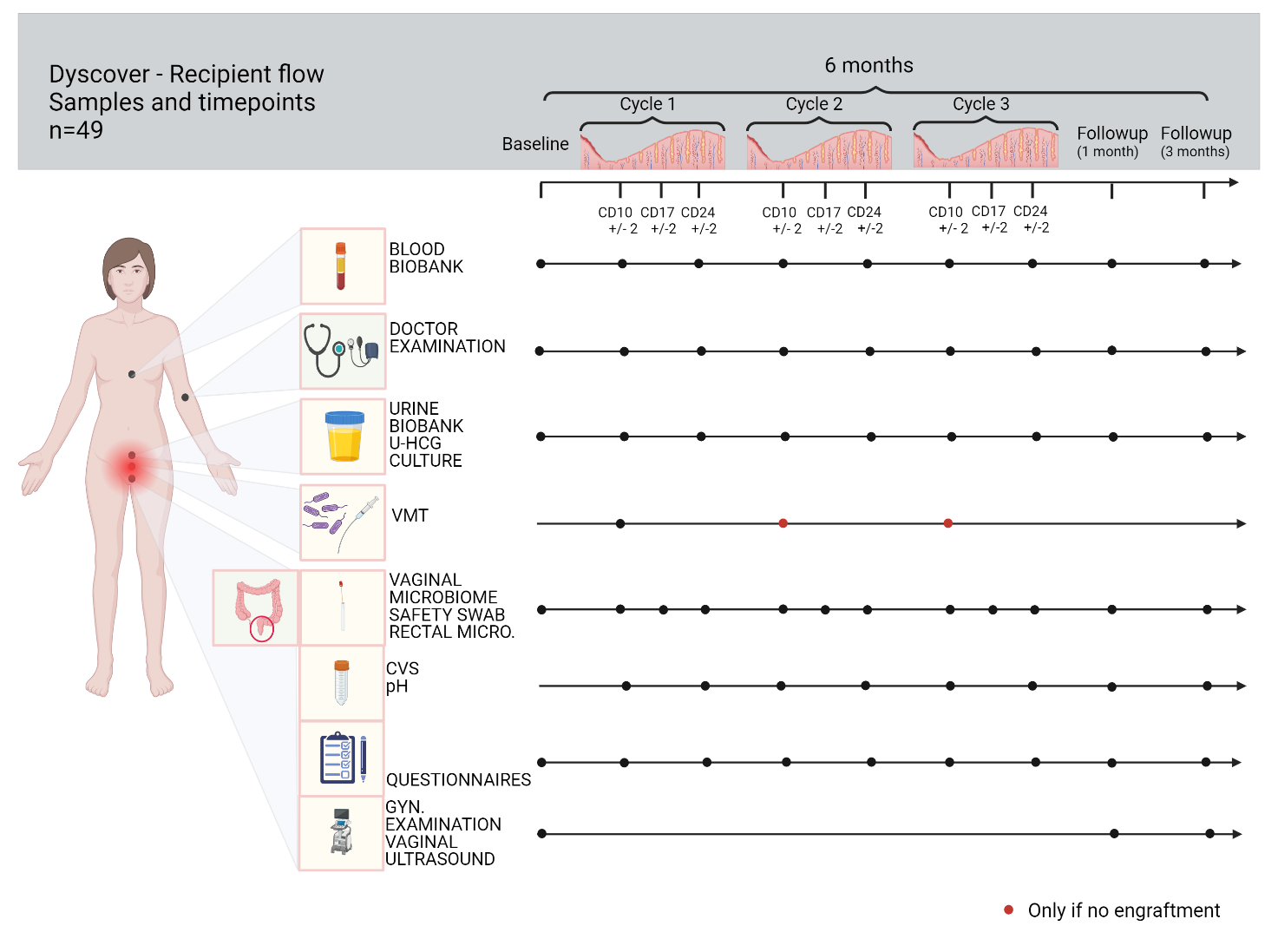

Figure 7

Study overview of participants and timeline of sample collections for women attending the flow cytometry study. Created with BioRender.com. CD, cycle day.

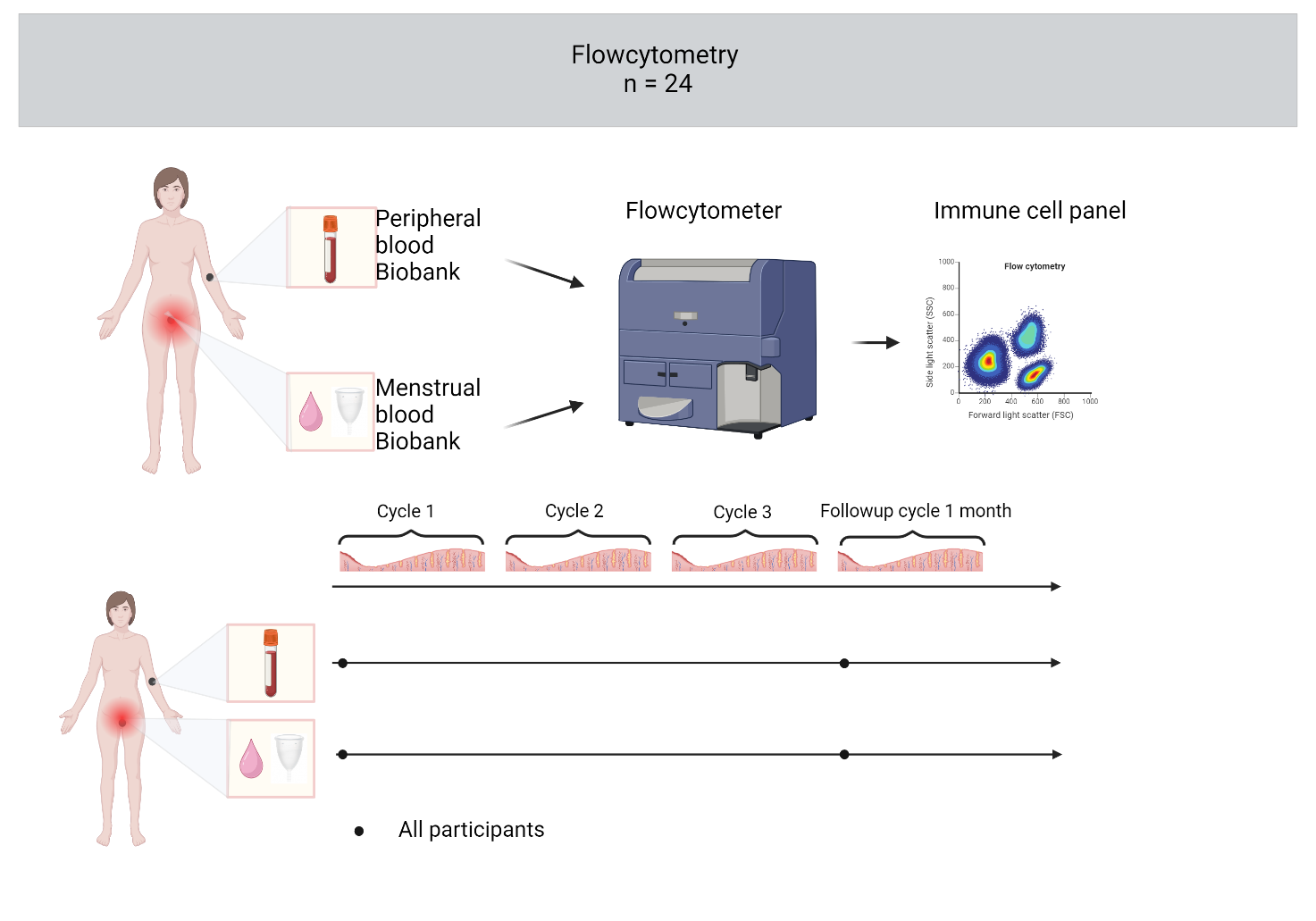

Figure 8

Study overview of participants and timeline of sample collections for women attending open-label extension part. Created with BioRender.com. CD, cycle day.

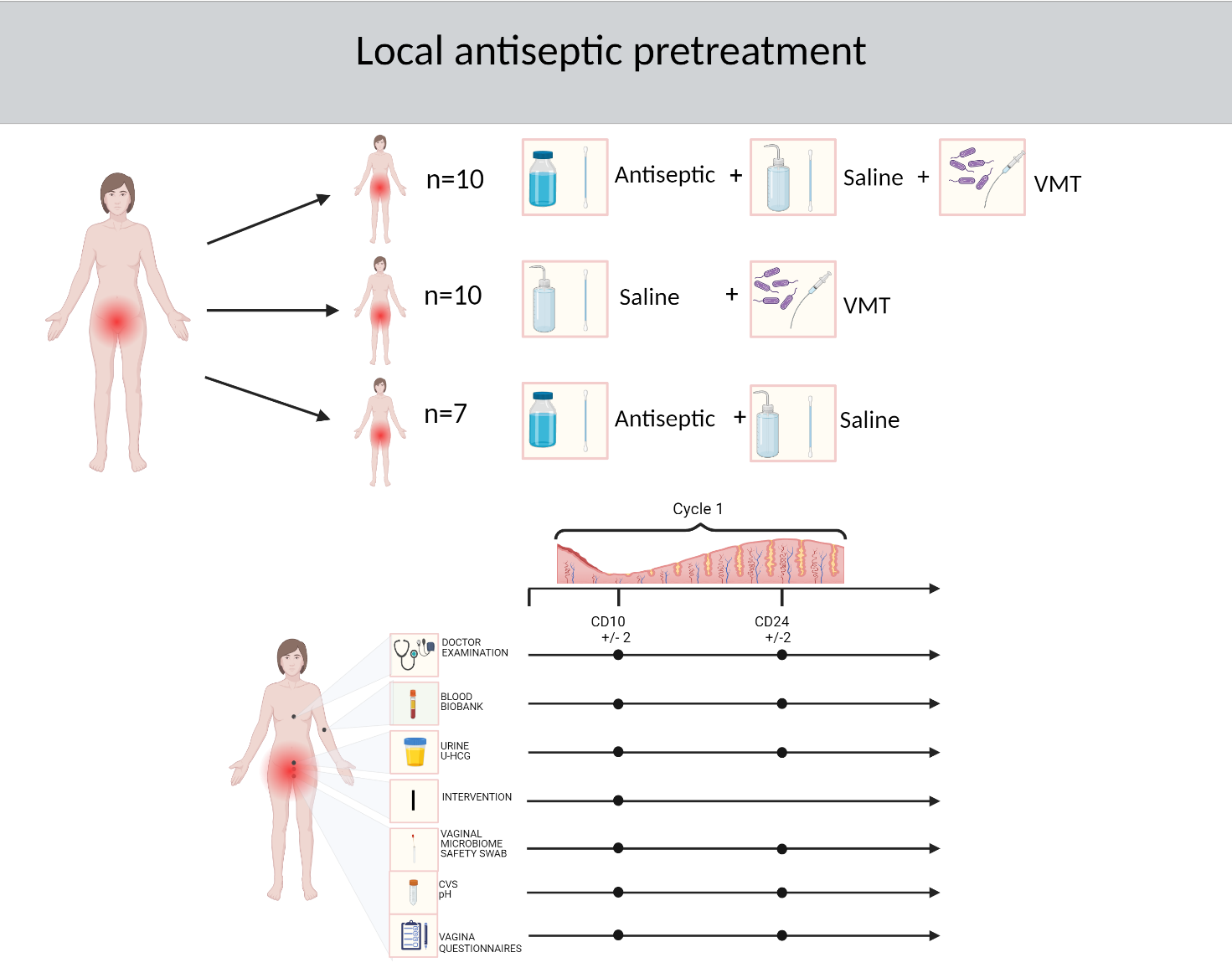
