## Supplementary tables for "Vaginal Microbiota Transplantation (VMT) for treatment of vaginal dysbiosis without the use of antibiotics – A Double-Blinded Randomized Controlled Trial in healthy women with vaginal dysbiosis"

Table 1

Overview of visits and samples for each visit in the donor group.

|  | **1.1** | **1.2** | **2.1** | **2.2** | **2.3** | **2.4** | **2.5** | **2.6** | **2.7** | **2.8** | **2.9** | **2.10** | **3** |
| --- | --- | --- | --- | --- | --- | --- | --- | --- | --- | --- | --- | --- | --- |
| **Study day** | **-40 til 0** | **0** | **1-31** | **2-32** | **3-33** | **4-34** | **5-35** | **6-36** | **7-37** | **8-38** | **9-39** | **10-40** | **28 -35 days after V2.10** |
| **Visit type** | **Screening** | **Screening** | **Donation** | **Donation** | **Donation** | **Donation** | **Donation** | **Donation** | **Donation** | **Donation** | **Donation** | **Donation** | **Followup** |
| **Informed consent** | **X** |  |  |  |  |  |  |  |  |  |  |  |  |
| **Eligibility for study** | **X** |  |  |  |  |  |  |  |  |  |  |  |  |
| **Review of medical history** | **X** |  |  |  |  |  |  |  |  |  |  |  |  |
| **Current medicine** | **X** |  |  |  |  |  |  |  |  |  |  |  |  |
| **Review of the questionnaire sent out** |  | **X** |  |  |  |  |  |  |  |  |  |  |  |
| **Submission of calendar** |  | **X** | **X** | **X** | **X** | **X** | **X** | **X** | **X** | **X** | **X** | **X** | **X** |
| **Demographic** |  | **X** |  |  |  |  |  |  |  |  |  |  |  |
| **Objective examination incl. vital values** |  | **X** |  |  |  |  |  |  |  |  |  |  | **X** |
| **Gynecological examination incl. ultrasound scan** |  | **X** |  |  |  |  |  |  |  |  |  |  | **X** |
| **Bloodsamples*** |  | **X** |  |  |  |  |  |  |  |  |  |  |  |
| **HIV 1/2** |  | **X** |  |  |  |  |  |  |  |  |  |  | **X** |
| **Hepatitis A, B, C** |  | **X** |  |  |  |  |  |  |  |  |  |  | **X** |
| **CMV** |  | **X** |  |  |  |  |  |  |  |  |  |  | **X** |
| **Treponema Palladum** |  | **X** |  |  |  |  |  |  |  |  |  |  | **X** |
| **Urine** |  |  |  |  |  |  |  |  |  |  |  |  |  |
| **Pregnantcy test** |  | **X** |  |  |  |  |  |  |  |  |  |  | **X** |
| **Urine analysis - culture** |  | **X** |  |  |  |  |  |  |  |  |  |  | **X** |
| **Vaginale swabs** |  |  |  |  |  |  |  |  |  |  |  |  |  |
| **Chlamydia trachomatis** |  | **X** |  |  |  |  |  |  |  |  |  |  | **X** |
| **Nesseria gonorrhoeae** |  | **X** |  |  |  |  |  |  |  |  |  |  | **X** |
| **Trichomonas vaginalis** |  | **X** |  |  |  |  |  |  |  |  |  |  | **X** |
| **Mycoplasma genitalium** |  | **X** |  |  |  |  |  |  |  |  |  |  | **X** |
| **Gr. A,B,C,G**  **Streptococci** |  | **X** |  |  |  |  |  |  |  |  |  |  | **X** |
| **Fungi** |  | **X** |  |  |  |  |  |  |  |  |  |  | **X** |
| Herpes Simplex (1+2) |  | **X** |  |  |  |  |  |  |  |  |  |  | **X** |
| **Human papilloma virus^#^** | **X** |  |  |  |  |  |  |  |  |  |  |  | **X** |
| **Mikrobiome analysis** | **X** |  | **X** | **X** | **X** | **X** | **X** | **X** | **X** | **X** | **X** | **X** |  |
| **CVS for analysis** |  |  |  |  |  |  |  |  |  |  |  |  |  |
| **Bacterial count** |  |  | **X** | **X** | **X** | **X** | **X** | **X** | **X** | **X** | **X** | **X** |  |
| **Seamen check** |  |  | **X** | **X** | **X** | **X** | **X** | **X** | **X** | **X** | **X** | **X** |  |
| **pH** |  |  | **X** | **X** | **X** | **X** | **X** | **X** | **X** | **X** | **X** | **X** |  |
| **CVS for VMT** |  |  | **X** | **X** | **X** | **X** | **X** | **X** | **X** | **X** | **X** | **X** |  |
| **Research Biobank** |  |  |  |  |  |  |  |  |  |  |  |  |  |
| **Blod** |  |  |  |  |  |  |  |  |  |  |  |  | **X** |
| **Urine** |  |  |  |  |  |  |  |  |  |  |  |  | **X** |
| **CVS** |  |  |  |  |  |  |  |  |  |  |  |  | **X** |
| **Vaginal swab**  **Rectal swab**  **Microbiome** | **X** |  |  |  |  |  |  |  |  |  |  |  | **X**  **X** |
| **Behavioral recommendations** |  | **X** | **X** | **X** | **X** | **X** | **X** | **X** | **X** | **X** | **X** | **X** |  |

Table 2

Overview of visits and samples for each visit in the recipients group for RCT study, flowcytometry and antiseptic pretreatment.

| **Cycle day** |  |  | **1** | **10** | **17** | **1** | **10** | **17** | **1** | **10** | **17** | **1** | **10** | **1** | **10** | **10** | **24** |
| --- | --- | --- | --- | --- | --- | --- | --- | --- | --- | --- | --- | --- | --- | --- | --- | --- | --- |
| **Visit** | **1.1** | **1.2** |  | **2** | **3** |  | **4** | **5** |  | **6** | **7** |  | **8** |  | **9** | **10** | **11** |
| **Study day** | **-28** | **-14** |  | **0** | **7** |  | **28** | **35** |  | **56** | **63** |  | **84** |  | **168** | **1 cykle after  V9** | **14 days after V10** |
| **Buffer in visits days** | **±14** | **±13** |  |  | **±2** |  | **±2** | **±2** |  | **±4** | **±4** |  | **±6** |  | **±12** | **±2** | **±2** |
| **Type of visit** | **Screening 1** | **Screening 2** | **Before BL** | **Baseline** | **Intervention** | **After 1. VMT**** | **Intervention** | **Intervention** | **After VMT with Engraftment**** | **Intervention** | **Follow-up** | **After VMT with Engraftment**** | **Follow-up** | **Menstrual blood collection follow -up** | **Follow-up** | **Intervention** | **Followup** |
| **Informed concent** | X |  |  |  |  |  |  |  |  |  |  |  |  |  |  | X |  |
| **Eligibility for study** | X |  |  |  |  |  |  |  |  |  |  |  |  |  |  |  |  |
| **Review of medical history** | X |  |  |  |  |  |  |  |  |  |  |  |  |  |  |  |  |
| **Current medicine** | X |  |  |  |  |  |  |  |  |  |  |  |  |  |  |  |  |
| **Review of the questionnaire sent out** |  | X |  |  |  |  |  |  |  |  |  |  |  |  |  |  |  |
| **Submission of calendar** |  | X |  | X | X |  | X | X |  | X | X |  | X |  | X |  |  |
| **Demographics** |  | X |  |  |  |  |  |  |  |  |  |  |  |  |  |  |  |
| **Sideeffects** |  |  |  |  | X |  | X | X |  | X | X |  | X |  | X |  |  |
| **Objective examination incl. vital values** |  | X |  | X | X |  | X | X |  | X | X |  | X |  | X | X | X |
| **Gynecological examination incl. ultrasound scan** |  | X |  |  | X |  | (X) | X |  | (X) | X |  | X |  | X |  |  |
| **Randomization** |  |  |  | X |  |  |  |  |  |  |  |  |  |  |  |  |  |
| **VMT procedure** |  |  |  | **X** |  |  | **(X)** |  |  | **(X)** |  |  |  |  |  | **X** |  |
| **Bloodsamples*** |  | X |  |  |  |  |  |  |  |  |  |  |  |  | X |  |  |
| **HIV 1/2** |  | X |  |  |  |  |  |  |  |  |  |  |  |  | X |  |  |
| **Hepatitis A,B,C** |  | X |  |  |  |  |  |  |  |  |  |  |  |  | X |  |  |
| **CMV** |  | X |  |  |  |  |  |  |  |  |  |  |  |  | X |  |  |
| **Treponema Palladum** |  | X |  |  |  |  |  |  |  |  |  |  |  |  | X |  |  |
| **Immuncells** |  |  | X |  |  |  |  |  | (X) |  |  | (X) |  | X |  |  |  |
| **Menstrual blood** |  |  | X |  |  |  |  |  | (X) |  |  | (X) |  | X |  |  |  |
| **Urin** |  |  |  |  |  |  |  |  |  |  |  |  |  |  |  |  |  |
| **Urine analysis - culture** |  | X |  | X |  |  |  |  |  |  |  |  |  |  | X |  | X |
| **Pregnancy** |  | X |  | X |  |  | X |  |  | X |  |  | X |  | X |  | X |
| **Vaginal swabs** |  |  |  |  |  |  |  |  |  |  |  |  |  |  |  |  |  |
| **Chlamydia trachomatis** |  | X |  | X |  |  | X |  |  | X |  |  | X |  | X |  | X |
| **Nesseria gonorrhoeae** |  | X |  | X |  |  | X |  |  | X |  |  | X |  | X |  | X |
| **Trichomonas vaginalis** |  | X |  | X |  |  | X |  |  | X |  |  | X |  | X |  | X |
| **Mycoplasma genitalium** |  | X |  | X |  |  | X |  |  | X |  |  | X |  | X |  | X |
| **Gr. A,B,C,G Streptococci** |  | X |  | X |  |  | X |  |  | X |  |  | X |  | X |  | X |
| **Herpes Simplex (1+2)** |  | X |  | X |  |  | X |  |  | X |  |  | X |  | X |  | X |
| **Fungi** |  | X |  | X |  |  | X |  |  | X |  |  | X |  | X |  | X |
| **Human papilloma virus^#^** |  | X |  |  |  |  |  |  |  |  |  |  |  |  | X |  | X |
| **Microbiome analysis** | X |  |  | X | X |  | X | X |  | X | X |  | X |  | X |  | X |
| **Research Biobank** |  |  |  |  |  |  |  |  |  |  |  |  |  |  |  |  |  |
| **Blood** |  |  |  | X | X |  | X | X |  | X | X |  | X |  | X | X | X |
| **Urine** |  |  |  | X | X |  | X | X |  | X | X |  | X |  | X |  |  |
| **CVS** |  |  |  | X | X |  | X | X |  | X | X |  | X |  | X | X | X |
| **Vaginal swab**  **Rectal swab**  **Microbiome** | X |  |  | X  X | X  X |  | X  X | X  X |  | X  X | X  X |  | X  X |  | X  X |  | X |
| **Behavioral recommendations** |  | X |  | X | X |  | X | X |  | X | X |  |  |  |  | X |  |

Table 3

Inclusion and exclusion criteria for donor and recipients

| Inclusion criteria |  |
| --- | --- |
| Donors | **Recipients** |
| Age 18-40 y/old | Age 18-40 y/old |
| Healthy and premenopausal | Healthy and premenopausal |
| Regular menstrual cycle and amenorrhea because of hormonal contraception | Regular menstrual cycle and amenorrhea because of hormonal contraception |
| Shotgun sequencing of vaginal swab showing normal, healthy microbiota | Shotgun sequencing of vaginal swab showing vaginal dysbiosis |
| Willing to answer personal questions regarding medical history and sexual health, behavior and history | Willing to answer personal questions regarding medical history and sexual health, behavior and history |
| Willing to follow behavioral restrictions during study participation | Willing to follow behavioral restrictions during study participation |
| Willing to collect cervicovaginal fluid up to 12 times | Willing to receive vaginal microbiota transplantation |
| No vaginal symptoms such as abnormal discharge, odor or itching |  |
| Exclusion criteria |  |
| Donors | **Recipients** |
| Pregnant or nursing | Pregnant, <8 weeks postpartum, nursing or planning a pregnancy in the next 6 months |
| Postmenopausal defined as >12 months amenorrhea with no known explanation | Postmenopausal defined as >12 months amenorrhea with no known explanation |
| IUD removal, cervical cryotherapy or laser treatment <3 months prior to study enrollment | IUD removal, cervical cryotherapy or laser treatment <3 months prior to study enrollment |
| Any condition requiring antibiotic treatment during study participation | Any condition requiring antibiotic treatment during study participation |
| Use of long-term hormonal treatments <3 months prior to study enrollment | Use of long-term hormonal treatments <3 months prior to study enrollment |
| Any condition, medical or psychiatric, assessed by the physician will make it unlikely for the participant to be able to follow the protocol | Any condition, medical or psychiatric, assessed by the physician will make it unlikely for the participant to be able to follow the protocol |
| Unsatisfying screening assessed by the examining physician | Unsatisfying screening assessed by the examining physician |
| Participation in other clinical trials <30 days prior to study enrollment | Participation in other clinical trials <30 days prior to study enrollment |
| Any history with BV, VD, treponema pallidum, M. genitalium, HPV including condylomas and CIN, herpes simplex 1+2, infections related to the pelvis, recurring UTI | Known infection with HIV, AIDS or other immune disease |
| N. gonorrhea or C. trachomatis <1 year prior to study participation |  |
| Positive HPV-test |  |
| Risk behavior assessed by the examining physician, including, use of medicine, travel and sexual behavior |  |
| Donors screened with positive CMV-IgG can be paired with CMV-IgG positive recipients |  |
| Hysterectomized |  |

Table 4

Overview of adverse events in relation to VMT material and total registration of AE’s in VMT and placebo group.

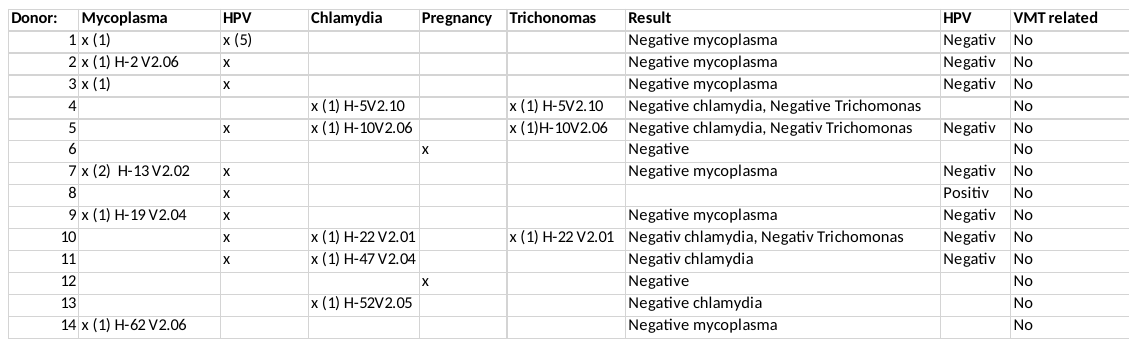

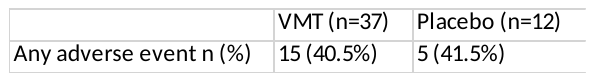
